# Does the habitual dietary intake of adults in Bavaria, Germany, match dietary intake recommendations? – Results of the 3^rd^ Bavarian Food Consumption Survey (BVS III)

**DOI:** 10.1101/2024.12.03.24318316

**Authors:** Florian Rohm, Nina Wawro, Sebastian Gimpfl, Nadine Ohlhaut, Melanie Senger, Christine Röger, Martin Kussmann, Kurt Gedrich, Jakob Linseisen

**Affiliations:** Chair of Epidemiology, University of Augsburg, Augsburg, Germany; Institute of Epidemiology, Helmholtz Centre Munich, Neuherberg, Germany; ZIEL – Institute for Food & Health, AG Public Health Nutrition, Technical University of Munich, Freising-Weihenstephan, Germany; Competence Center for Nutrition (KErn), Bavarian Research Institution for Agriculture (LfL), Freising, Germany; Institute for Medical Information Processing, Biometry, and Epidemiology, Medical Faculty, Ludwig-Maximilians University Munich, Munich, Germany

**Keywords:** Dietary intake, Bavaria, BVS III, NCI method, 24-hour-recalls, Nutrients, Food (groups), Food-based dietary guidelines (FBDGs)

## Abstract

1

**Objective:** Monitoring dietary habits is crucial for identifying shortcomings and delineating countermeasures. About 20 years after the last population-based surveys in Bavaria and Germany, dietary habits were assessed to describe the intake distributions and compare these with recommendations at food and nutrient level.

**Methods:** The 3^rd^ Bavarian Food Consumption Survey (BVS III) was designed as a diet survey representative of adults in Bavaria; from 2021 to 2023, repeated 24-hour diet recalls were collected by telephone using the software GloboDiet©. Food (sub-)group and nutrient intake data were modeled with the so-called NCI method, weighted for the deviation from the underlying population. Intake distributions in men and women were described as percentiles. These data were used to estimate the proportion of persons meeting dietary intake recommendations. In addition, food consumption data were compared with the results reported 20 years ago collected by the same methodology (2^nd^ Bavarian Food Consumption Survey, BVS II).

**Results:** Using 24-hour diet recalls of 550 male and 698 female participants, we estimated intake distributions for food (sub-)groups and nutrients. A major proportion of the adult population does not meet the food-based dietary guidelines; this refers to a series of food groups, including fruit and vegetables, legumes, nuts, cereal products, and especially whole grain products, as well as fresh and processed meat. Regarding selected essential nutrients, a considerable proportion of the population was at higher risk of insufficiency from iron (women), zinc (men), and folic acid (both men and women), as already described in previous studies.

**Conclusion:** A major proportion of the adult Bavarian population does not meet the current food-based dietary guidelines. Compared to BVS II data, favorable changes refer to lower consumption of total meat (especially processed meat) and soft drinks, and an increased intake of vegetables. The conclusions based on the intake of selected essential nutrients hardly changed over time. From a public health perspective, the still low intake of vegetables, fruit, nuts, cereal products, and particularly of whole grain products, and associated higher risks of insufficient supply of several vitamins and minerals call for action for improvement.

## 2 Introduction

Since the 2^nd^ Bavarian Food Consumption Survey (BVS II, 2002–2003) and the National Food Consumption Survey II (NVS II, 2005–2007) with four follow-up surveys in the National Nutrition Monitoring (NEMONIT), no population-based surveys in adults with a direct recording of dietary intake were conducted in Germany or any federal state of Germany. The 3^rd^ Bavarian Food Consumption Survey (BVS III) aimed to close this gap for Bavaria and to provide current cross-sectional data on food consumption and nutrient intake of the adult population in Bavaria.

Food consumption survey methods are designed to estimate the dietary intake of a defined population. When the dietary intake distribution of a population is estimated based on a single-day measurement, the intake distribution contains between-person information while the within-person variation is not captured. This means that the variance of the usual group intake is inflated by day-to-day variation in individual intake. Repeated 24-hour diet recalls (24HR) allow to account for this intra-individual variability. Several statistical methods were developed over the past decades to estimate usual intake distributions from repeated 24HR, taking into account intra-personal variation (e.g., (1–7)). The approach developed at the National Cancer Institute NCI, commonly referred to as NCI method (6, 7), allows the inclusion of covariates when modeling intake distributions. The inclusion of a food frequency questionnaire (FFQ) as a covariate and thus combining two measurement instruments is of particular interest (8). Using the BVS II study data, we previously investigated the differences in food intake distributions by comparing the results of a weighted means approach and the NCI method (9). The estimation of valid intake distributions is a necessary precondition for evaluating the percentage of the population meeting intake recommendations.

In 2024, the German Society for Nutrition (DGE) published the results of a mathematical optimization model for deriving food-based recommendations (10, 11). So far, these values have not been evaluated using population-based intake data. Therefore, this study aimed at estimating the most valid food and nutrient intake distributions for the adult Bavarian population and describing the agreement with reference values.

## 3 Methods

### 3.1 Study population

The BVS III was planned as a representative study for the Bavarian population aged 18 to 75 years. In a two-stage random procedure (random selection of municipalities and random selection of subjects within these municipalities via the residents’ registration offices), potential study participants were contacted. After removing quality-neutral non-participants, 1,503 men and women aged 18 to 75 years were surveyed, i.e., 26% of the persons in the gross sample.

### 3.2 Recruitment and data collection

The household visits took place in the time frame of October 2021 to November 2022, and the nutrition survey was conducted until January 2023. Thus, the entire study framework lay within the period of the SARS-CoV-2 pandemic.

During the face-to-face interview in the households, information on sociodemographic characteristics, diet-related behavior (including a short food frequency questionnaire (FFQ) covering approximately 30 foods and food groups), and on the health status of the participants was assessed. Additionally, participants completed self-administered questionnaires per tablet, e.g., on physical activity.

Dietary intake data were collected by 24HR during the six weeks following the home visit. Per subject, three 24HR should be completed on randomly selected days (two weekdays, one weekend day). To ensure standardized assessment, the software GloboDiet©, a further development of the EPIC-SOFT© software, which was used in the BVS II (12), was applied. The 24HR were conducted as computer-assisted telephone interviews (CATI) by trained interviewers. Subsequently, the data underwent intensive quality control. From 1,239 persons, one (n=91), two (n=165), or three (n=983) 24HR were available and used for the statistical analysis.

All individual food items in the 24HR were assigned a code according to the German food composition database (Bundeslebensmittelschlüssel, BLS) (13), version 3.02. The foods were aggregated into main food groups and subgroups based on the hierarchical BLS coding system. In addition, the subgroups “fermented milk products”, the main food group “alternative products” with its subgroups “milk alternatives” and “meat alternatives”, as well as the main food group “whole grain products” were newly defined. Additionally, we defined the following food groups: “total meat” (sum of fresh meat and processed meat), “red meat” (fresh meat minus poultry), “fruit and vegetables” (sum of fruit and vegetables), and “cereal products” (sum of bread and bakery products, staple food, and whole grain products). Dairy consumption was converted into milk equivalents according to Breidenassel et al. (14).

### 3.3 Covariates

Self-reported body weight and height were used to calculate Body Mass Index (BMI; kg/m²). BMI subgroups were established according to the WHO definition (15). Smoking was described as never, ex-, or current smokers. Habitual physical activity was assessed employing the validated EHIS-PAQ (16). Each person’s physical activity level was described with one of the following categories: sedentary, low active, active, or very active (17).

Based on their information on their highest school and professional qualification according to the International Standard Classification of Education (ISCED), the participants were assigned the corresponding ISCED 97 level (18). According to the ISCED classification of the Federal Statistical Office and the German Microdata Lab, the assigned ISCED 97 levels were grouped into 3 educational levels (19): low educational group (levels 1 and 2), medium educational group (levels 3 and 4), and high educational group (levels 5 and 6).

The net equivalent income was calculated using information on net household income and household composition. For this purpose, the corresponding average value was first assigned to each income group queried (e.g., 1,250 euros for “1,000 to less than 1,500 euros”). Household size was weighted using the weighting factors of the modified Organization for Economic Co-operation and Development (OECD) scales (20). The first adult is weighted with a factor of 1.0, while the other household members aged 14 and over are weighted with a factor of 0.5, and all others with 0.3. The net equivalized income of the participants was calculated by dividing the net household income by the weighted household size. Classification into low, medium, and high income was carried out along the lines of risk of poverty and income wealth (21). A net equivalent income below 60% of the national median income is considered low, while net equivalent income above 200% of the national median income is considered high. The median national equivalized income in Germany in 2022, when most of the data collection in the BVS III took place, was 25,000 euros/year (22).

### 3.4 Description of Weighting

To ensure representativeness for the Bavarian population, the nutritional data was weighted, based on the 2020 micro-census and intercensal population updates for Bavaria as a reference. The weighting was conducted to correct for the oversampling of the Augsburg study area and non-response, considering administrative district, political municipality size class, education level, gender, and age.

### 3.5 Statistical Analysis

The descriptive analysis of characteristics of the study population was conducted separately for men and women. Results are given as arithmetic means and standard deviation or absolute and relative frequencies, as appropriate.

The NCI method (6, 7) was applied to estimate the distribution of habitual food and nutrient intakes separately for men and women. The NCI method is based on the idea that the usual intake can be understood as the probability of consumption multiplied by the amount consumed. The approach follows a two-step procedure by estimating the consumption probability of a food item by a logistic regression and the amount of consumption of a food item by a linear model separately. Both parts can be linked by allowing for a correlation of the person-specific effects included in the models. In both models, age, gender, BMI and education level were included as covariates. If available, FFQ information was also included as a covariate in the probability model. Additionally, a population-weighting variable was specified and for each 24HR, the information on whether it was recorded on a weekday or a weekend day was included. Intake estimates of daily consumed food items and nutrients were derived without fitting the probability model. For these calculations, the SAS macros MIXTRAN V2.1 and DISTRIB V2.1 provided by the National Cancer Institute (NCI) of the National Institute of Health (NIH) were used.

Statistical analysis was performed using SAS software, version 9.4 of the SAS System for Windows (Copyright © 2002–2010 SAS Institute Inc.).

Habitual dietary intake estimates were compared with recommendations published by the DGE. To evaluate food group intake data, the newly released food-based dietary guidelines (FBDG), precisely the results of the „optimization model 2“, were used (10, 11). Habitual vitamin and mineral intakes were compared with the most recent reference values published by the German Nutrition Society, except for retinol equivalents (23).

## 4 Results

### 4.1 Characteristics of the sample population

Results from the descriptive (not weighted) analysis are presented in Table 1. In the present study, 550 male (44%) and 689 female (56%) participants with a mean age of 48.6 and 49.2 years, respectively, were analyzed. The obesity prevalence was 21.6% and 15.1% in male and female participants, respectively. More than 62% of men were pre-obese or obese, while the corresponding figure in women was 43%. The proportion of current smokers was lower in women (14%) as compared to men (19%). About 31% of women and 22% of men followed a sedentary level of physical activity. The proportion of very active subjects was about twice as high in men as in women (31% versus 17%). About half of the participants had a high education and roughly 20% a low education; based on their self-reports, 14% were classified as having a high net equivalence income, while 26% (males) and 28% (females) were attributed to the low-income group.

**Table 1:**
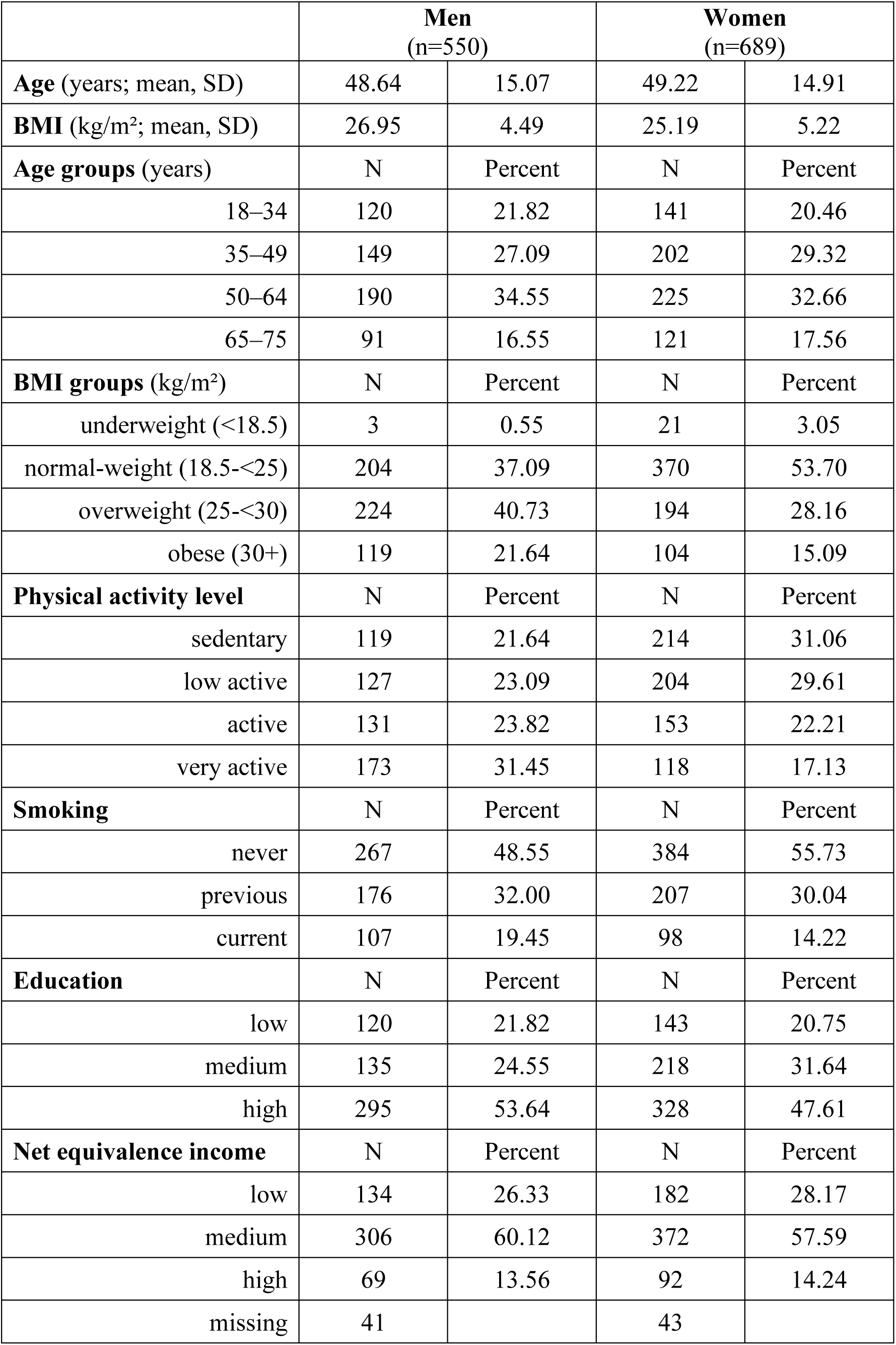
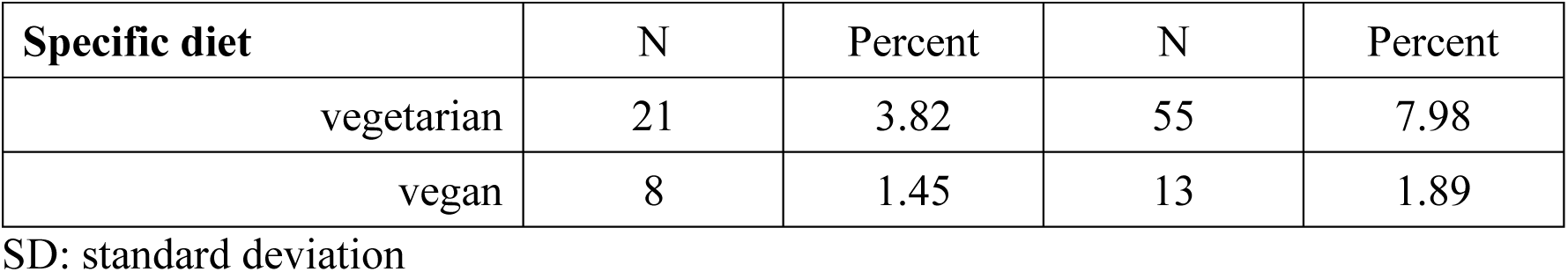
Characteristics of male and female participants of the BVS III.

|  | **Men**<br>(n=550) |  | **Women**<br>(n=689) |  |
| --- | --- | --- | --- | --- |
| **Age** (years; mean, SD) | 48.64 | 15.07 | 49.22 | 14.91 |
| **BMI** (kg/m<sup>2</sup>; mean, SD) | 26.95 | 4.49 | 25.19 | 5.22 |
| **Age groups** (years) | N | Percent | N | Percent |
| 18–34 | 120 | 21.82 | 141 | 20.46 |
| 35–49 | 149 | 27.09 | 202 | 29.32 |
| 50–64 | 190 | 34.55 | 225 | 32.66 |
| 65–75 | 91 | 16.55 | 121 | 17.56 |
| **BMI groups** (kg/m<sup>2</sup>) | N | Percent | N | Percent |
| underweight (<18.5) | 3 | 0.55 | 21 | 3.05 |
| normal-weight (18.5-<25) | 204 | 37.09 | 370 | 53.70 |
| overweight (25-<30) | 224 | 40.73 | 194 | 28.16 |
| obese (30+) | 119 | 21.64 | 104 | 15.09 |
| **Physical activity level** | N | Percent | N | Percent |
| sedentary | 119 | 21.64 | 214 | 31.06 |
| low active | 127 | 23.09 | 204 | 29.61 |
| active | 131 | 23.82 | 153 | 22.21 |
| very active | 173 | 31.45 | 118 | 17.13 |
| **Smoking** | N | Percent | N | Percent |
| never | 267 | 48.55 | 384 | 55.73 |
| previous | 176 | 32.00 | 207 | 30.04 |
| current | 107 | 19.45 | 98 | 14.22 |
| **Education** | N | Percent | N | Percent |
| low | 120 | 21.82 | 143 | 20.75 |
| medium | 135 | 24.55 | 218 | 31.64 |
| high | 295 | 53.64 | 328 | 47.61 |
| **Net equivalence income** | N | Percent | N | Percent |
| low | 134 | 26.33 | 182 | 28.17 |
| medium | 306 | 60.12 | 372 | 57.59 |
| high | 69 | 13.56 | 92 | 14.24 |
| missing | 41 |  | 43 |  |

| Specific diet | N | Percent | N | Percent |
| --- | --- | --- | --- | --- |
| vegetarian | 21 | 3.82 | 55 | 7.98 |
| vegan | 8 | 1.45 | 13 | 1.89 |

SD: standard deviation

### 4.2 Habitual food consumption

Data on food consumption in men and women are provided in Tables 2 and 3, and in the supplementary Tables S1 and S2.

**Table 2:**
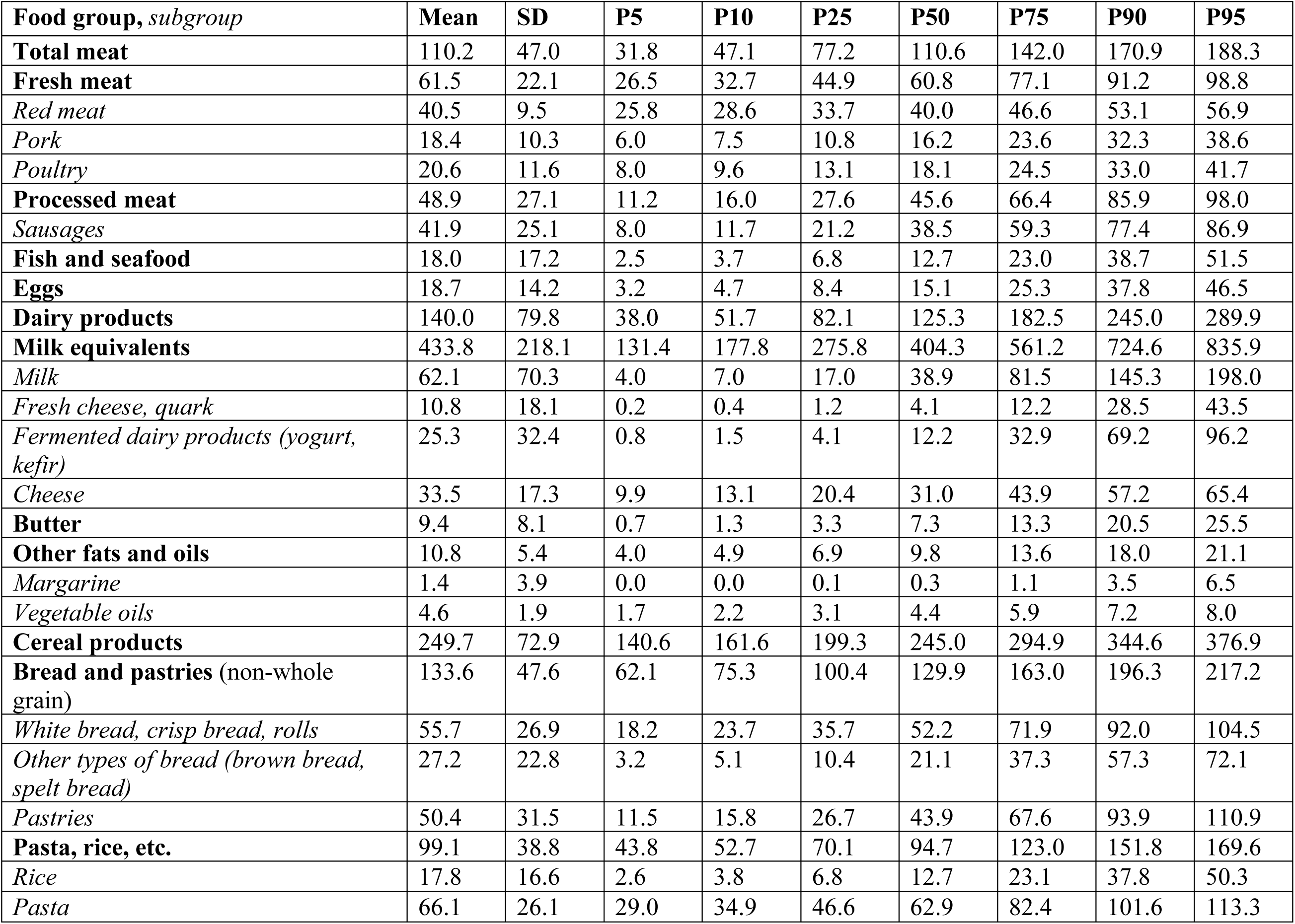
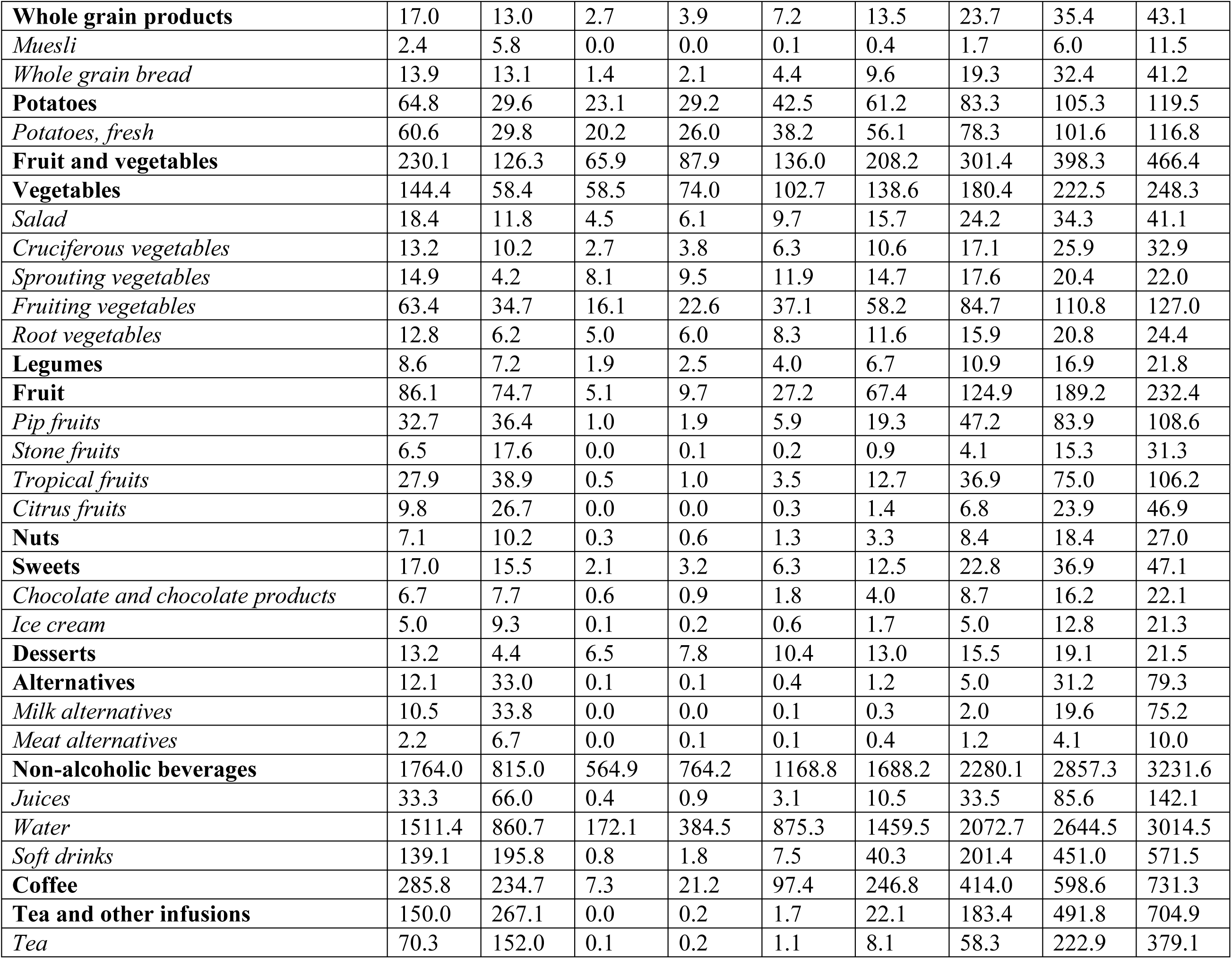
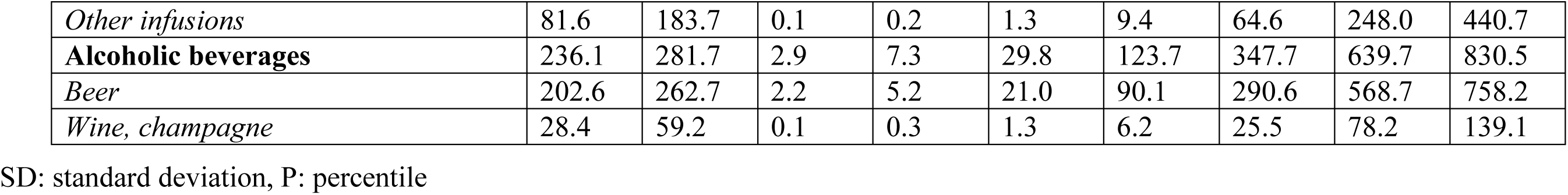
Food group consumption distribution (g/day) in male participants (n=550) of the BVS III, weighted for the deviation from the underlying population.

**Table 3:**
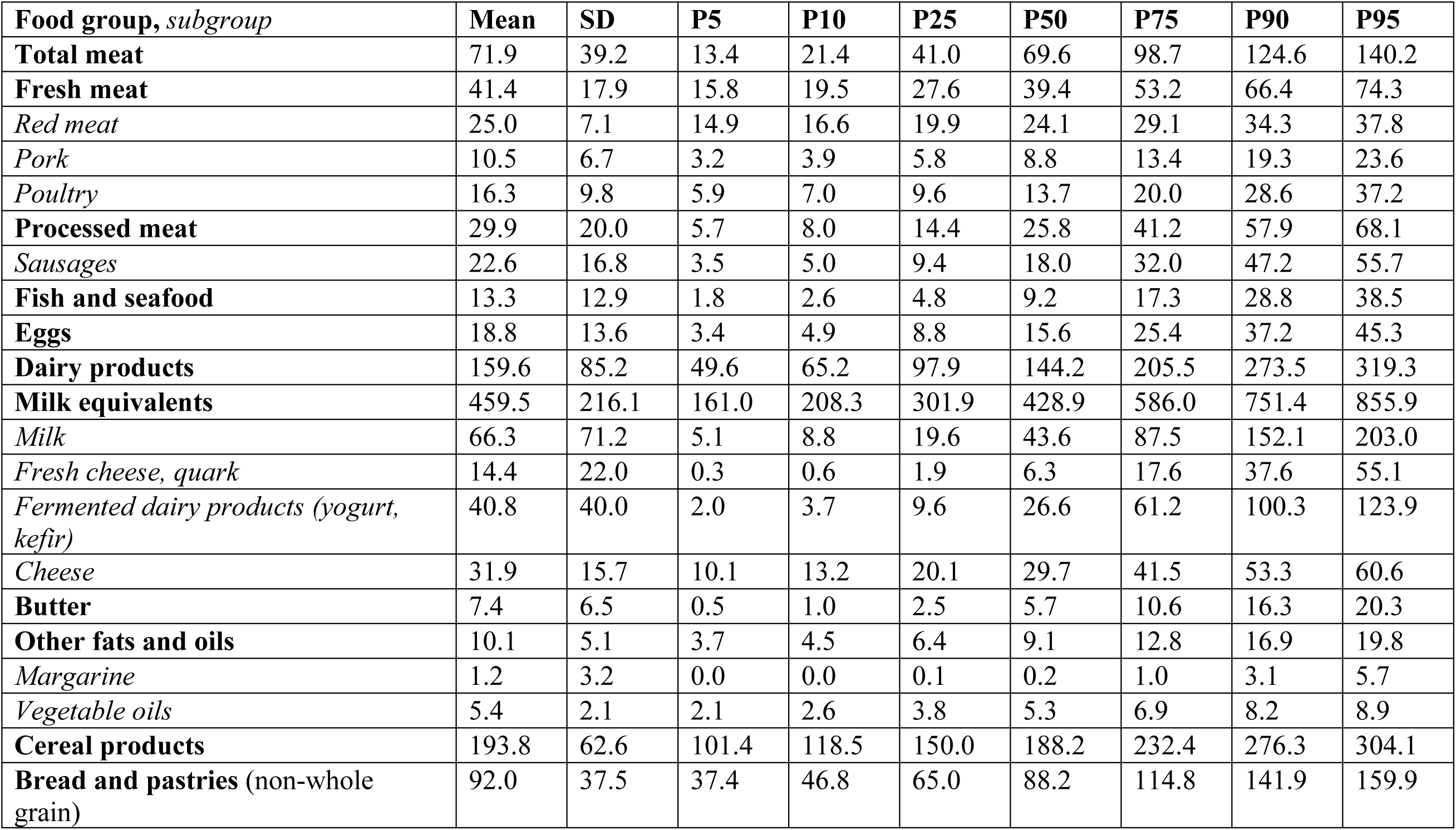
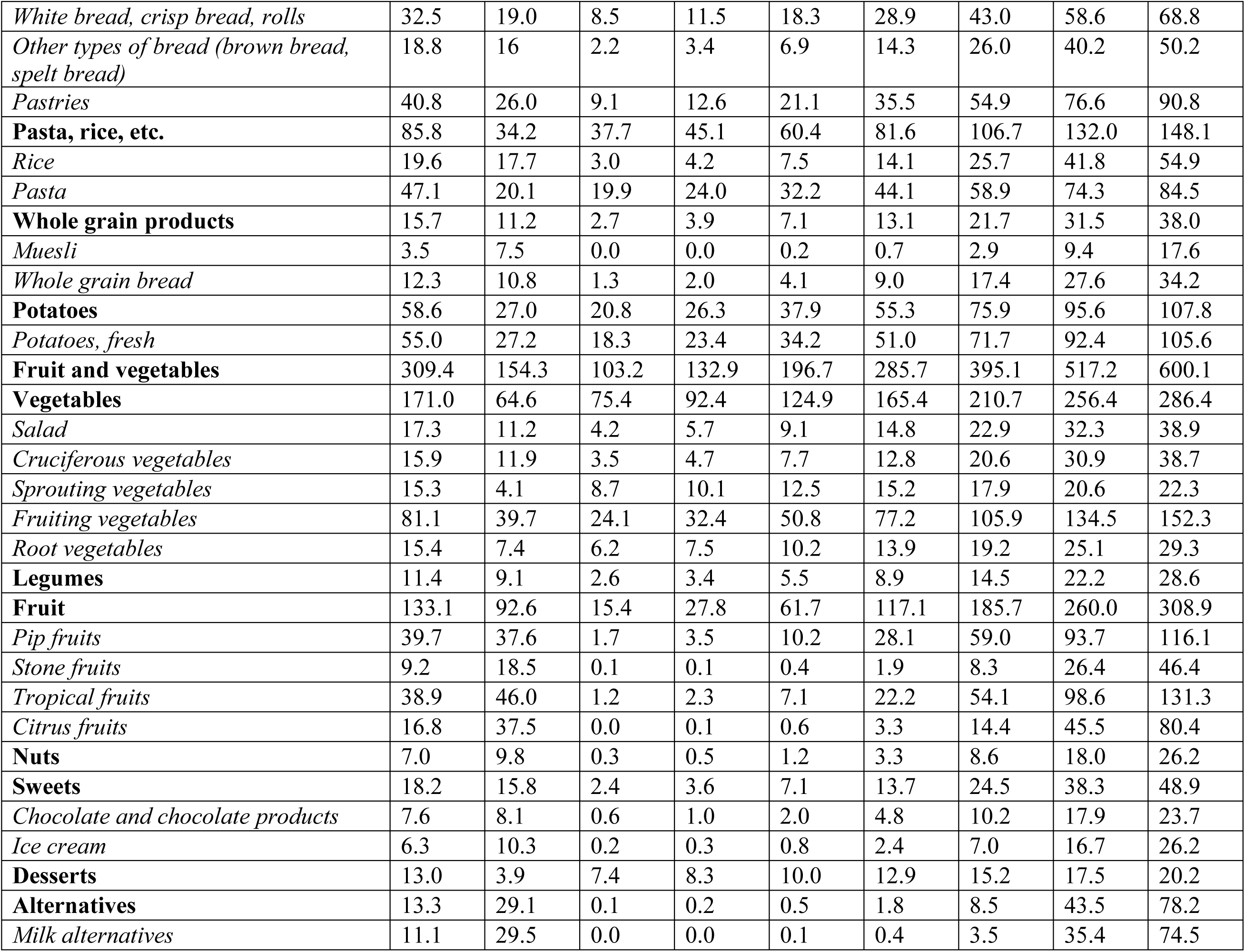
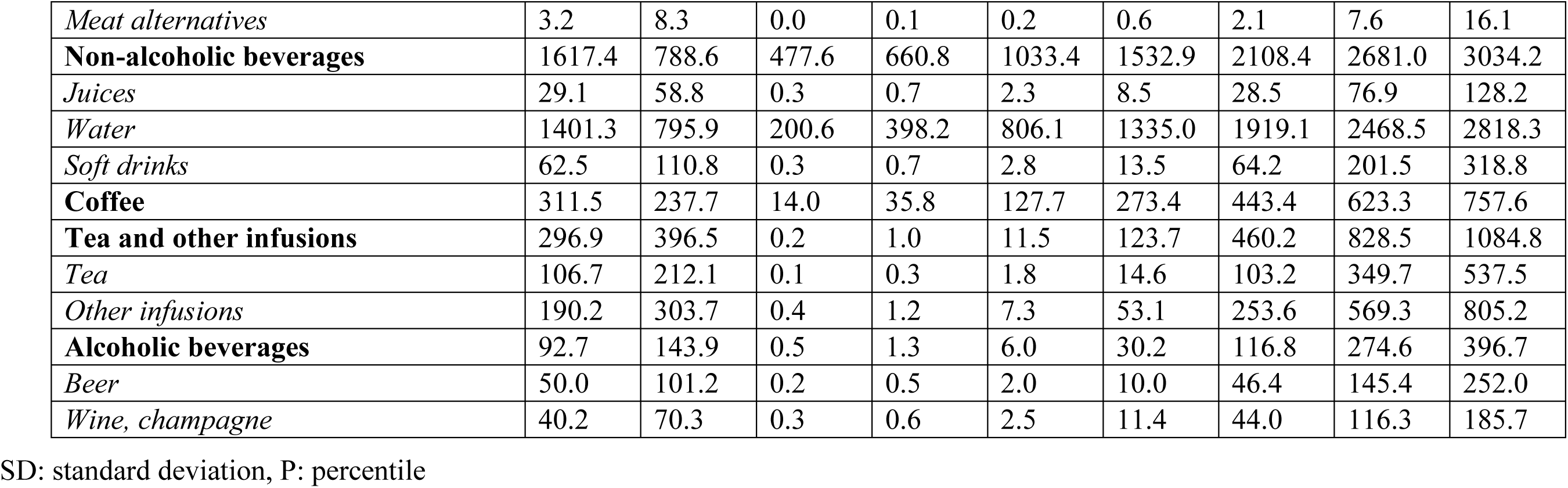
Food group consumption distributions (g/day) in female participants (n=689) of the BVS III, weighted for the deviation from the underlying population.

The median (25th–75th percentile) consumption of fresh meat was 60.8 (44.9–77.1) g/day for men and 39.4 (27.6–53.2) g/day for women; additionally, 45.6 (27.6–66.4) g/day and 25.8 (14.4–41.2) g/day of processed meat were consumed, respectively. Arithmetic means were generally higher, indicating skewed distributions. The Bavarian diet is low in fish and seafood, with median consumption figures of 12.7 (6.8–23.0) g/day and 9.2 (4.8–9.2) g/day. Median egg consumption was about 15 g/day in both sexes. Women consumed more fermented milk products (yogurt, kefir) than men; when expressed in milk equivalents (MEq), men consumed 404 (276–561) g/day and women 429 (302–586) g MEq/day.

The median intake of meat and milk alternatives was low (1.2 and 1.8 g/day in males and females, respectively), i.e., half of the population consumed hardly any alternatives. 25% of the population (75th percentile) consumed at least 5.0 and 8.5 g/day, and 10% (90th percentile) consumed 31.2 and 43.5 g/day or more. Consumption of milk alternatives (about 80% of the alternatives) dominated over meat alternatives (about 20%).

Among fats and oils, butter and vegetable oils were the major contributors, while median margarine intake was very low (0.3 g/day). The median consumption of butter was highest with 7.3 (3.3–13.3) g/day in men and 5.7 (2.5–10.6) g/day in women. Median consumption of vegetable oils amounted to 4.4 (3.1–5.9) g/day in men and 5.3 (3.8–6.9) g/day in women.

The median (25th–75th percentile) daily consumption of vegetables amounted to 138.6 (102.7– 180.4) g/day for men and 165.4 (124.9–210.7) g/day for women. Also, daily fruit consumption was distinctly lower in men with 67.4 (27.2–124.9) g compared to women with 117.1 (61.7–185.7) g. The median daily consumption of potatoes amounted to 56 and 55 g in men and women, respectively. Median consumption of nuts was low with 3.3 g/day both in men and women. Median consumption of cereal products amounted to 245.0 g/day in men and 188.2 g/day in women. Major contributors were bread and pasta.

The dominating subgroup among non-alcoholic beverages was water (1.5 (0.9–2.1) l/day in men and 1.3 (0.8–1.3) l/day in women), followed by coffee (247 (97–414) ml/day in men and 273 (128–443) ml/day in women). Consumption of soft drinks was higher in men with 40.3 (7.5–201.4) ml/day compared to women with 13.5 (2.8–64.2) ml/day in women.

Men drank more alcoholic beverages, especially beer, than women. Median intake data for beer was 90.1 ml/d in men and 10.0 ml/d in women; for wine, median intake data were 6.2 ml/d in men and 11.4 ml/d in women. Mean values were distinctly higher indicating substantially skewed distributions.

The comparison of these intake data with the German food-based dietary guidelines (Table 4) shows that the recommendations on the consumption of plant-derived food, including fruit and vegetables, nuts, whole grain products, and vegetable oils were only met by a minor proportion of the population (<16%). Exceptions are only the food groups potatoes and legumes. On the contrary, red and processed meat, whose intakes exceed the FBDG for at least 88% of the population, are consumed in higher amounts than recommended. Median consumption of dairy products is slightly above the recommended amounts, with 47% of the men and 43% of the women consuming less than the corresponding FBDG.

**Table 4:**
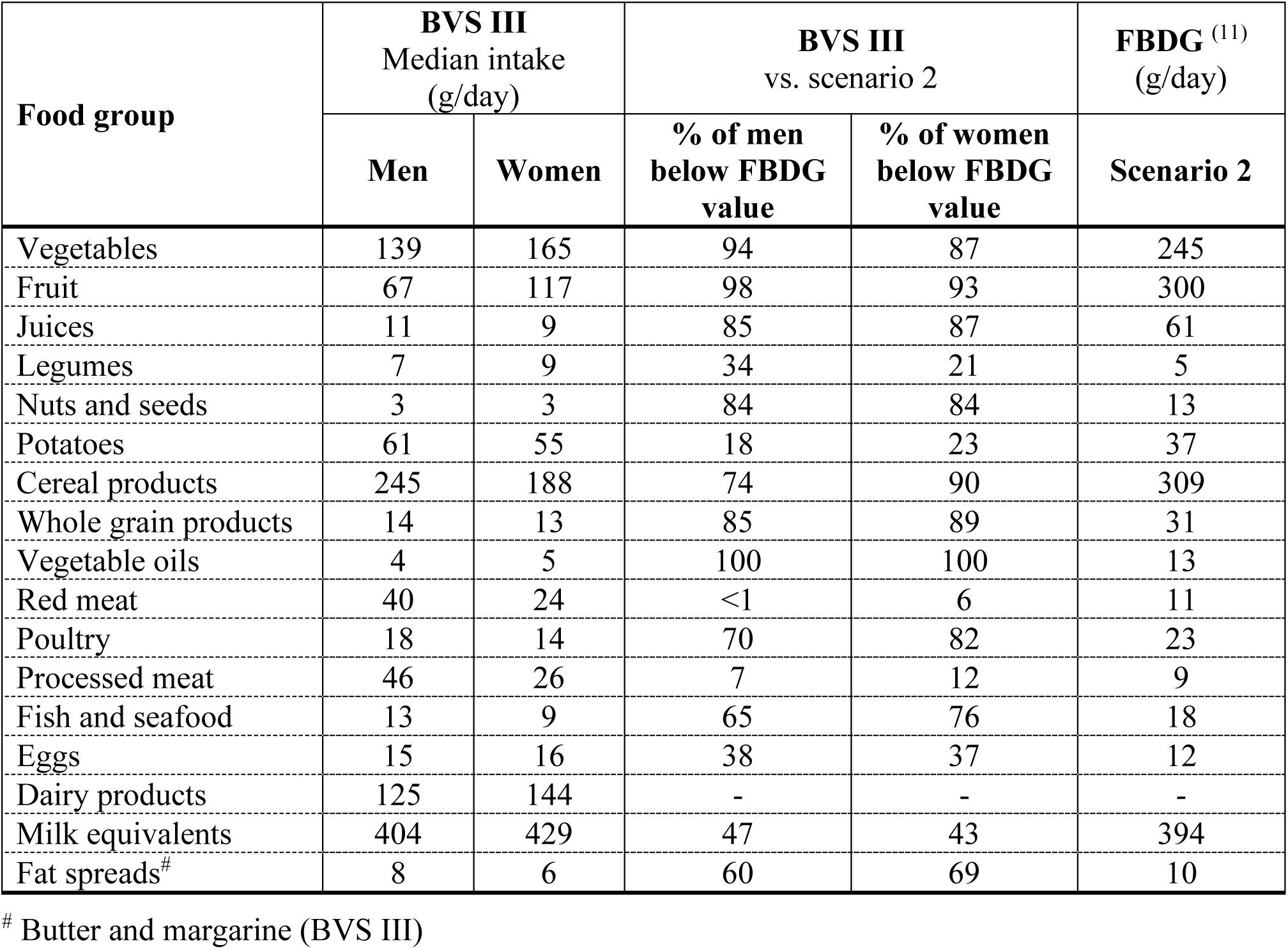
Median intake and proportion of male and female participants of the BVS III meeting the food-based dietary guidelines (scenario 2) of the German Nutrition Society (10, 11)

| Food group | BVS III<br>Median intake<br>(g/day) |  | BVS III<br>vs. scenario 2 |  | FBDG <sup>(11)</sup><br>(g/day) |
| --- | --- | --- | --- | --- | --- |
|  | Men | Women | % of men<br>below FBDG<br>value | % of women<br>below FBDG<br>value | Scenario 2 |
| Vegetables | 139 | 165 | 94 | 87 | 245 |
| Fruit | 67 | 117 | 98 | 93 | 300 |
| Juices | 11 | 9 | 85 | 87 | 61 |
| Legumes | 7 | 9 | 34 | 21 | 5 |
| Nuts and seeds | 3 | 3 | 84 | 84 | 13 |
| Potatoes | 61 | 55 | 18 | 23 | 37 |
| Cereal products | 245 | 188 | 74 | 90 | 309 |
| Whole grain products | 14 | 13 | 85 | 89 | 31 |
| Vegetable oils | 4 | 5 | 100 | 100 | 13 |
| Red meat | 40 | 24 | <1 | 6 | 11 |
| Poultry | 18 | 14 | 70 | 82 | 23 |
| Processed meat | 46 | 26 | 7 | 12 | 9 |
| Fish and seafood | 13 | 9 | 65 | 76 | 18 |
| Eggs | 15 | 16 | 38 | 37 | 12 |
| Dairy products | 125 | 144 | - | - | - |
| Milk equivalents | 404 | 429 | 47 | 43 | 394 |
| Fat spreads<sup>#</sup> | 8 | 6 | 60 | 69 | 10 |

<sup>#</sup> Butter and margarine (BVS III)

Compared to the results of the BVS II, the median intake of vegetables increased, more distinctly in women than in men, but the median fruit intake remained stable in women and decreased in men (Table 5). A distinct difference was noted for processed meat consumption; men and women lowered their median intake by 40–48% compared to the amount reported 20 years ago. Also, the median intake of red meat slightly decreased. The same is true for fish and dairy products. Median poultry and egg consumption increased. Regarding beverages, a much higher median water consumption was noted, while beer (in men) and wine consumption (in men and women) decreased. In the case of skewed distributions and high intakes in less than 50% of the population, median values do not reflect changes in this subgroup.

**Table 5:** Median consumption of food groups in male and female participants of the BVS III compared to the BVS II.

| Food group | BVS III (2021-23)<br>(g/day) |  | BVS II (2002–03) <sup>(9)</sup><br>(g/day) |  |
| --- | --- | --- | --- | --- |
|  | Men | Women | Men | Women |
| Vegetables | 139 | 165 | 127 | 129 |
| Fruit | 67 | 117 | 80 | 116 |
| Nuts | 3 | 3 | 1 | 1 |
| Potatoes | 61 | 55 | 63 | 58 |
| Cereal products | 245 | 188 | 237 | 175 |
| Vegetable oils | 4 | 5 | 4 | 4 |
| Margarine | 0.3 | 0.2 | 1 | 1 |
| Butter | 7 | 6 | 10 | 8 |
| Red meat | 40 | 24 | 48 | 30 |
| Poultry | 18 | 14 | 15 | 12 |
| Processed meat | 46 | 26 | 87 | 43 |
| Fish and seafood | 13 | 9 | 18 | 14 |
| Eggs | 15 | 16 | 9 | 8 |
| Dairy products | 125 | 144 | 146 | 165 |
| Water | 1460 | 1335 | 613 | 768 |
| Soft drinks | 40 | 14 | 31 | 11 |
| Beer | 90 | 10 | 242 | 10 |
| Wine | 6 | 11 | 24 | 19 |

### 4.3 Habitual consumption of energy and nutrients

The habitual consumption of energy and nutrients is shown in Table 6 (for males) and Table 7 (for females). The median (25th–75th percentile) daily energy intake was 1974 (1688–2283) kcal/day in men and 1588 (1338–1858) kcal/day in women. The median intake of saturated fatty acids and the sum of mono- and disaccharides were 35.6 g/day and 74.1 g/day in men, and 23.8 g/day and 51.1 g/day in women, respectively. Median dietary fiber intake in men and women was about 16 g/day, and men consumed twice the amount of ethanol than women (median intake of 15.7 g/day in men and 7.8 g/day in women).

**Table 6:**
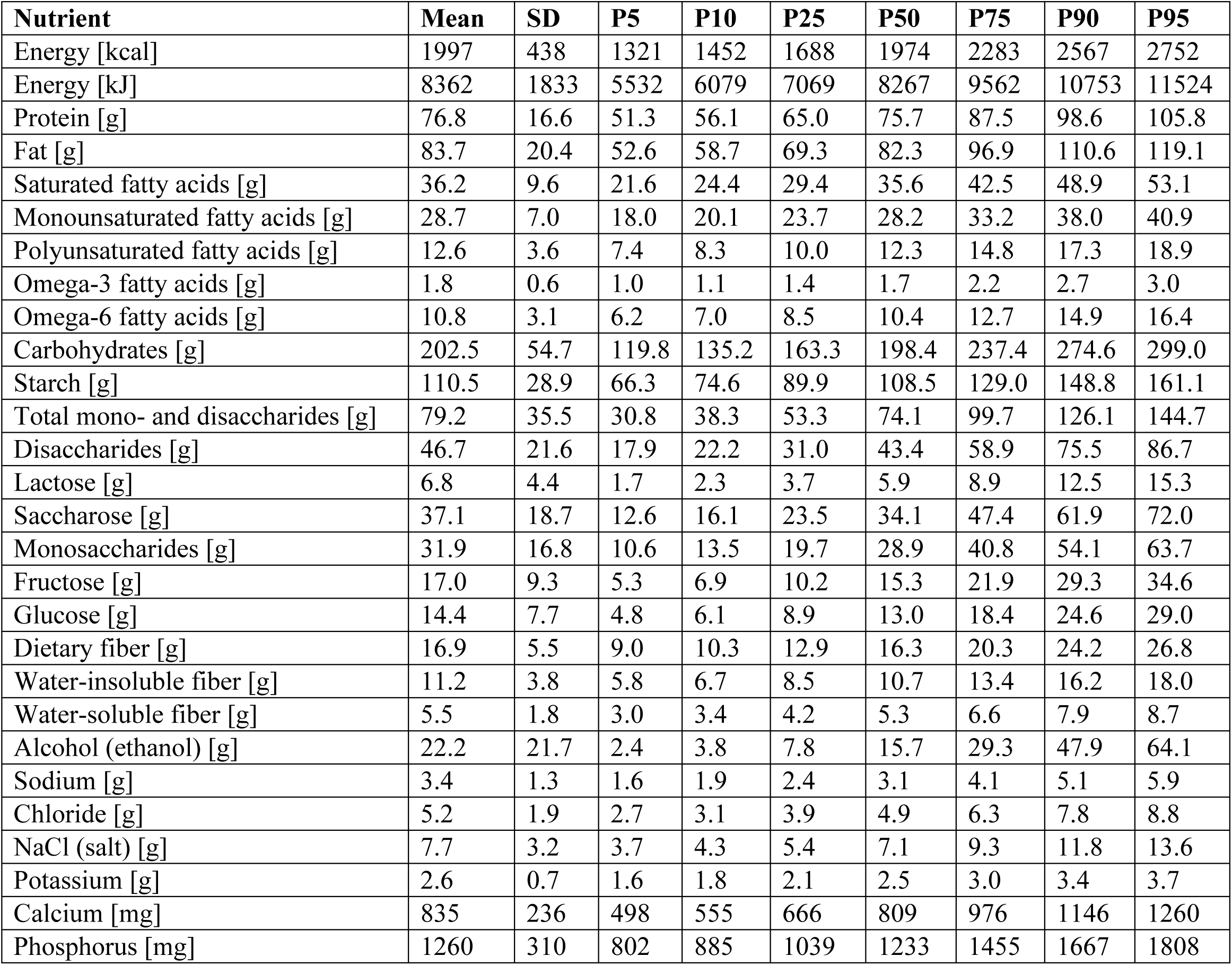
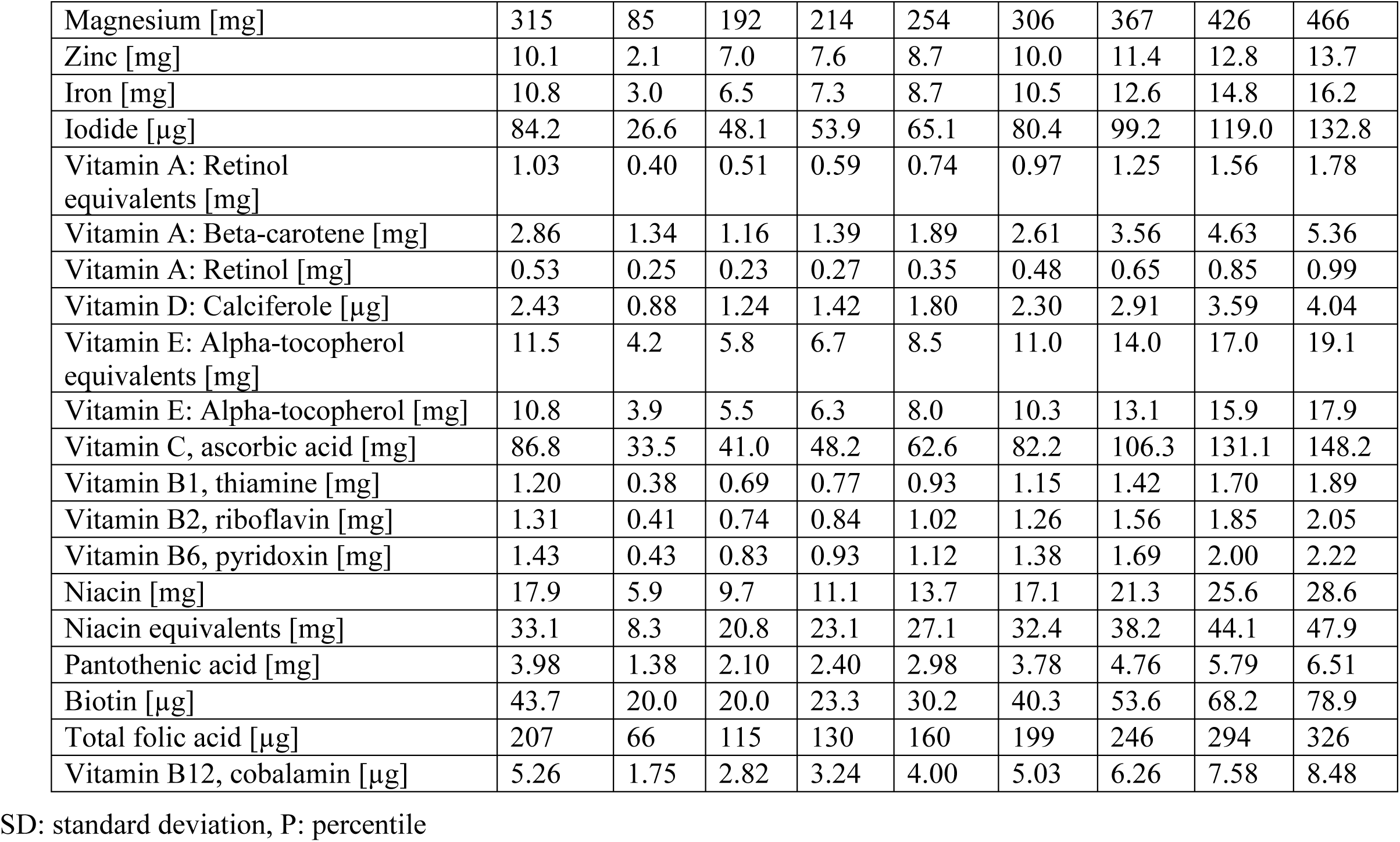
Energy and nutrient intake distributions (g/day) in male participants (n=550) of the BVS III, weighted for the deviation from the underlying population.

**Table 7:** Energy and nutrient intake distributions (g/day) in female participants (n=689) of the BVS III, weighted for the deviation from the underlying population.

| Nutrient | Mean | SD | P5 | P10 | P25 | P50 | P75 | P90 | P95 |
| --- | --- | --- | --- | --- | --- | --- | --- | --- | --- |
| Energy [kcal] | 1612 | 393 | 1011 | 1128 | 1338 | 1588 | 1858 | 2132 | 2303 |
| Energy [kJ] | 6753 | 1646 | 4235 | 4723 | 5603 | 6649 | 7782 | 8929 | 9644 |
| Protein [g] | 60.0 | 14.5 | 38.1 | 42.4 | 49.7 | 59.0 | 69.0 | 79.0 | 85.8 |
| Fat [g] | 70.4 | 18.5 | 42.4 | 47.8 | 57.4 | 69.1 | 81.9 | 95.0 | 103.2 |
| Saturated fatty acids [g] | 30.0 | 8.7 | 16.9 | 19.4 | 23.8 | 29.3 | 35.4 | 41.6 | 45.5 |
| Monounsaturated fatty acids [g] | 23.8 | 6.3 | 14.4 | 16.1 | 19.3 | 23.3 | 27.7 | 32.1 | 35.0 |
| Polyunsaturated fatty acids [g] | 11.2 | 3.3 | 6.5 | 7.3 | 8.8 | 10.8 | 13.1 | 15.5 | 17.1 |
| Omega-3 fatty acids [g] | 1.6 | 0.6 | 0.8 | 1.0 | 1.2 | 1.5 | 1.9 | 2.4 | 2.7 |
| Omega-6 fatty acids [g] | 9.5 | 2.9 | 5.4 | 6.1 | 7.5 | 9.2 | 11.2 | 13.3 | 14.7 |
| Carbohydrates [g] | 165.2 | 49.3 | 92.0 | 105.5 | 130.3 | 160.8 | 195.6 | 230.8 | 253.1 |
| Starch [g] | 85.9 | 25.7 | 47.1 | 54.4 | 67.4 | 83.8 | 102.0 | 119.9 | 131.4 |
| Total mono- and disaccharides [g] | 76.6 | 35.2 | 29.0 | 36.4 | 51.1 | 71.4 | 96.2 | 123.9 | 142.1 |
| Disaccharides [g] | 44.7 | 21.3 | 16.6 | 20.8 | 29.3 | 41.2 | 56.1 | 73.2 | 84.5 |
| Lactose [g] | 7.6 | 4.7 | 2.0 | 2.7 | 4.3 | 6.6 | 9.9 | 13.8 | 16.7 |
| Saccharose [g] | 36.7 | 18.9 | 12.1 | 15.7 | 22.9 | 33.5 | 46.8 | 62.0 | 72.3 |
| Monosaccharides [g] | 32.6 | 17.3 | 10.7 | 13.7 | 20.0 | 29.3 | 41.4 | 55.7 | 65.5 |
| Fructose [g] | 17.7 | 9.8 | 5.5 | 7.1 | 10.6 | 15.8 | 22.6 | 30.6 | 36.3 |
| Glucose [g] | 14.5 | 7.8 | 4.8 | 6.1 | 8.8 | 13.0 | 18.4 | 24.9 | 29.3 |
| Dietary fiber [g] | 16.2 | 5.4 | 8.4 | 9.7 | 12.3 | 15.6 | 19.3 | 23.3 | 26.0 |
| Water-insoluble fiber [g] | 10.8 | 3.7 | 5.5 | 6.4 | 8.1 | 10.4 | 12.9 | 15.7 | 17.6 |
| Water-soluble fiber [g] | 5.1 | 1.7 | 2.7 | 3.1 | 3.9 | 4.9 | 6.0 | 7.3 | 8.1 |
| Alcohol (ethanol) [g] | 12.1 | 13.4 | 0.9 | 1.6 | 3.6 | 7.8 | 15.5 | 27.5 | 37.5 |
| Sodium [g] | 2.4 | 1.0 | 1.2 | 1.3 | 1.7 | 2.2 | 2.9 | 3.6 | 4.2 |
| Chloride [g] | 4.0 | 1.5 | 2.0 | 2.3 | 2.9 | 3.7 | 4.7 | 5.9 | 6.7 |
| NaCl (salt) [g] | 5.5 | 2.3 | 2.6 | 3.0 | 3.8 | 5.0 | 6.6 | 8.4 | 9.8 |
| Potassium [g] | 2.4 | 0.6 | 1.4 | 1.6 | 1.9 | 2.3 | 2.8 | 3.2 | 3.5 |
| Calcium [mg] | 788 | 227 | 464 | 521 | 625 | 762 | 922 | 1087 | 1202 |
| Phosphorus [mg] | 1035 | 269 | 642 | 713 | 845 | 1009 | 1196 | 1391 | 1519 |
| Magnesium [mg] | 279 | 77 | 168 | 188 | 224 | 270 | 324 | 381 | 417 |
| Zinc [mg] | 8.3 | 1.8 | 5.5 | 6.1 | 7.0 | 8.2 | 9.4 | 10.7 | 11.5 |
| Iron [mg] | 9.6 | 2.7 | 5.7 | 6.4 | 7.6 | 9.3 | 11.1 | 13.2 | 14.5 |
| Iodide [µg] | 74.3 | 23.6 | 42.4 | 47.6 | 57.5 | 70.8 | 87.1 | 105.5 | 118.2 |
| Vitamin A: Retinol equivalents [mg] | 1.10 | 0.43 | 0.54 | 0.62 | 0.79 | 1.03 | 1.32 | 1.66 | 1.90 |
| Vitamin A: Beta-carotene [mg] | 3.46 | 1.59 | 1.43 | 1.73 | 2.32 | 3.17 | 4.28 | 5.52 | 6.48 |
| Vitamin A: Retinol [mg] | 0.50 | 0.24 | 0.21 | 0.25 | 0.34 | 0.45 | 0.61 | 0.81 | 0.95 |
| Vitamin D: Calciferole [µg] | 2.32 | 0.84 | 1.18 | 1.36 | 1.71 | 2.19 | 2.79 | 3.44 | 3.90 |
| Vitamin E: Alpha-tocopherol equivalents [mg] | 11.1 | 4.1 | 5.6 | 6.5 | 8.2 | 10.6 | 13.4 | 16.5 | 18.7 |
| Vitamin E: Alpha-tocopherol [mg] | 10.3 | 3.7 | 5.2 | 6.0 | 7.6 | 9.7 | 12.3 | 15.2 | 17.2 |
| Vitamin C, ascorbic acid [mg] | 99.1 | 37.3 | 47.9 | 56.1 | 72.4 | 94.0 | 120.0 | 149.0 | 168.1 |
| Vitamin B1, thiamine [mg] | 0.99 | 0.31 | 0.57 | 0.63 | 0.77 | 0.94 | 1.16 | 1.40 | 1.57 |
| Vitamin B2, riboflavin [mg] | 1.13 | 0.36 | 0.63 | 0.71 | 0.88 | 1.09 | 1.34 | 1.62 | 1.81 |
| Vitamin B6, pyridoxin [mg] | 1.16 | 0.36 | 0.66 | 0.74 | 0.90 | 1.11 | 1.36 | 1.63 | 1.82 |
| Niacin [mg] | 13.9 | 4.8 | 7.3 | 8.4 | 10.5 | 13.2 | 16.5 | 20.2 | 22.7 |
| Niacin equivalents [mg] | 25.7 | 7.0 | 15.6 | 17.4 | 20.8 | 25.0 | 29.9 | 34.9 | 38.2 |
| Pantothenic acid [mg] | 3.58 | 1.26 | 1.88 | 2.15 | 2.68 | 3.39 | 4.27 | 5.25 | 5.93 |
| Biotin [μg] | 40.8 | 20.0 | 18.6 | 21.7 | 28.2 | 37.5 | 49.6 | 64.1 | 74.4 |
| Total folic acid [μg] | 197 | 64 | 108 | 122 | 151 | 189 | 233 | 281 | 314 |
| Vitamin B12, cobalamin [μg] | 3.92 | 1.41 | 2.01 | 2.31 | 2.91 | 3.71 | 4.70 | 5.77 | 6.54 |

SD: standard deviation, P: percentile

When compared with the reference values of the DGE, the proportion of persons below these values is lowest for retinol equivalents, vitamin B2, niacin, and vitamin B12 (Table 8). A high proportion of the population not meeting the reference values was identified for folic acid, pantothenic acid, and vitamin B6, i.e., a substantial proportion of the population was at higher risk of insufficient supply of these nutrients. This also applies to the habitual intake of iodine, potassium, calcium, magnesium, zinc in men, and iron, especially in premenopausal women, for which up to 100% of the population did not meet the reference values.

**Table 8:** Median intake of selected vitamins and proportion of male and female participants of the BVS III not meeting the reference values of the German Nutrition Society (DGE)

| Vitamin | Daily intake, BVS III (median) |  | % of participants below the reference value |  | Reference value (DGE<sup>1</sup>) |  |
| --- | --- | --- | --- | --- | --- | --- |
|  | Men | Women | Men | Women | Men | Women |
| Vitamin A, Retinol equivalents [mg] | 0.97 | 1.03 | 25 | 11 | 0.75 | 0.65 |
| Vitamin E, α-tocopherol equivalents [mg] | 11.0 | 10.6 | 75 | 63 | 14 | 12 |
| Vitamin B1 [mg] | 1.15 | 0.94 | 55 | 57 | 1.2 | 1.0 |
| Vitamin B2 [mg] | 1.26 | 1.09 | 63 | 39 | 1.4 | 1.1 |
| Vitamin B6 [mg] | 1.38 | 1.11 | 69 | 78 | 1.6 | 1.4 |
| Niacin equivalents [mg] | 32.4 | 25.0 | 0 | 0 | 15 | 12 |
| Pantothenic acid [mg] | 3.78 | 3.39 | 79 | 87 | 5 | 5 |
| Folic acid, total [μg] | 199 | 189 | 91 | 93 | 300 | 300 |
| Vitamin B12 [μg] | 5.03 | 3.71 | 24 | 58 | 4 | 4 |
| Vitamin C [mg] | 82.2 | 94.0 | 77 | 51 | 110 | 95 |

<sup>1</sup> Reference values for 25–50-year-old men and women (33), except for retinol equivalents (23)

## 5 Discussion

### 5.1 Summary of findings

In this population-based study, current data on the intake of food groups and nutrients are presented for men and women. The precise intake distribution was modeled using the NCI method. The results provided information on dietary changes over the past 20 years and their (dis-)agreement with food-based dietary guidelines as recently released in Germany. The most pronounced and favorable changes refer to a lower consumption of processed meat (including sausages) and beer (in men). Median intake of vegetables increased especially in women. However, in many aspects, the observed diet deviates substantially from the respective recommendations and guidelines, e.g., on fruit and vegetable intake. Unfavorably low intakes of whole grain products and fruit and vegetables on the one hand, and high intakes of red and processed meat on the other hand are still prevalent. Vitamin and mineral intake result from food selection and no improvement over the past shortcomings was observed.

### 5.2 Methodological aspects

In this population-based study, 26% of the eligible persons participated eventually. Notably, the prevailing SARS-CoV-2 pandemic was an important confounding factor. After the home visit, 1,239 persons completed 24HR. To account for biases from differential non-response, all analyses were weighted for the deviation from the underlying population.

Misreporting, especially underreporting, is a persistent problem in dietary assessment, leading to an underestimation of dietary intake (24). Obesity being a major determinant of the likelihood of underreporting (24), and the prevalence of obesity being comparable in both BVS II and BVS III, the extent of the problem of misreporting seems fairly stable. In addition, we used the same method for dietary assessment in both studies, i.e., telephone interviews conducted by trained interviewers using the same software, to ensure a highly comparable and standardized protocol.

Due to the limited number of participants, the NCI method did not allow for the estimation of intake distributions for further subgroups, e.g., age groups. Another limitation of the NCI method is that it cannot identify non-consumers. Therefore, estimated population distributions do not enable the identification of the proportion of the population not consuming a certain food item.

### 5.3 Evaluation of habitual food consumption

The proportion of persons following a vegetarian or vegan diet in adult Bavarians increased over the past years (12); at the same time, people also followed the concept of a flexitarian diet, i.e., limiting the number of days with meat-based dishes. As the median meat consumption decreased, the observation of decreasing meat consumption is not driven by the group of vegetarians and vegans but rather reflects a broad change in eating behavior in the population.

Although the consumption of red meat and especially processed meat has decreased among men and women over the past 20 years (Table 5), current consumption levels reported here are on average higher than the recommended levels of the FBDG; in men, less than 1% met the FBDG for red meat, and 7% were in line with the FBDG for processed meat.

Median dairy product consumption decreased by about 20 g/day over the past decades (BVS II). When converted into milk equivalents (14), the intakes of 43% of men and 47% of women are below the FBDG.

The comparison of the consumption of foods of plant origin with the FBDG revealed significant deviations (Table 4). Adult men and women in Bavaria consumed far fewer vegetables and fruit in 2021–2023 than recommended, with 10% or less of men and women meeting the fruit and vegetable intake recommendations. To a similar extent, this also applies to the consumption of nuts and seeds, with more than 80% of the adult population eating fewer nuts and seeds than recommended. However, vegetable intake increased over the past 20 years, especially in women.

The median potato intake on the other hand exceeded the corresponding FBDG by almost 90%, while the consumption of cereal products was distinctly lower than recommended. Moreover, the FBDG for whole grain products were missed by far: 85% of all men and almost 90% of all women did not meet the recommendations. In the BVS II, similar amounts of potatoes were consumed on average (Table 5), and the consumption of cereal products increased slightly since then.

For the first time, the consumption of milk alternatives as well as meat alternatives is reported in the BVS III and indicates an increasing importance of milk alternatives in particular. These findings are supported by market data: In the past years, the consumption of alternatives for dairy and meat products has grown continuously, although the absolute contribution is still rather low, with dairy product alternatives making up 6.6% of the total dairy market in 2023 in Germany (25).

A positive development is the distinct decrease in the consumption of soft drinks, beer, and wine in the BVS III compared to 20 years ago, while at the same time, the median consumption of drinking water has almost doubled (Table 5).

Major observed dietary changes (compared to BVS II), especially the decreased meat consumption, were mirrored by data from food balance sheets (26).

### 5.4 Evaluation of habitual consumption of vitamins and minerals

The modeling of the nutrient intake distribution, correcting for intrapersonal variation, enables the evaluation of micronutrient intakes by identifying the proportion of the Bavarian population with an intake below or above reference values. We used the reference values of the German Nutrition Society (11) established to ensure that almost all persons of the population met their nutrient requirements when meeting these values. Accordingly, men and women not meeting these reference values have a higher risk of insufficient intake of the respective nutrients; for diagnosing nutrient deficiency, biochemical analyses of biomarkers in biospecimens are warranted. German reference data for average requirements in the population were not established but would represent the preferred concept for comparison.

The largest proportion of persons below reference values was observed for folic acid, exceeding 90% in both sexes and corresponding to previous findings in Germany (27). However, available biomarker data to evaluate the supply status of folic acid in the German population described the problem precisely (28). In addition, high proportions of individuals not meeting recommendations were also observed for pantothenic acid, vitamin E, and vitamin B6. For all three vitamins, lower risks of insufficiency were observed in previous studies in Germany (28, 29). The high proportion of men (more than 75%) not reaching the reference values for vitamin C (110 mg/d) was particularly surprising, which may be explained by the low median intake of vegetables and particularly of fruit in men (Table 4). The proportion of men and women below the DGE reference values for vitamins was lowest for niacin; also, for retinol equivalents and vitamin B12 (in men), low proportions were observed. We did not include vitamin D in this comparison because diet usually constitutes only a minor contribution to vitamin D supply (30). Overall, we did not observe distinct differences to former studies as summarized by Bechthold et al. (31, 32).

In terms of mineral intakes, low proportions of the population not meeting the DGE reference values were observed for phosphorus, sodium, and chloride (Table 9). It should be noted, however, that the intake of sodium and chloride cannot be precisely assessed using 24HD, since, e.g., adding salt (NaCl) at consumption is not recorded, resulting in an underestimation of the intake of these minerals. On the other hand, large parts of the population not meeting the reference values were observed for iodine, calcium, magnesium, zinc in men, and iron in women (both pre- and postmenopausal). To determine the actual iron supply status, established biomarker measurements would have to be performed. Similar observations concerning these minerals were made for Germany in the NVS II, yet to a lesser extent (29). Surprisingly, also the median potassium intake was 40% lower than the DGE reference value, resulting in 97 and 98% of men and women, respectively, not meeting the reference values. Possible reasons may include the inadequate consumption of potassium-rich foods, particularly fruit, vegetables, nuts, and cereal products (Table 4), as well as underreporting.

**Table 9:** Median intake of selected minerals and proportion of male and female participants of the BVS III not meeting the reference values of the German Nutrition Society (DGE)

| Mineral | Daily intake, BVS III (median) |  | % of participants below the reference value |  | Reference value (DGE<sup>1</sup>) |  |
| --- | --- | --- | --- | --- | --- | --- |
|  | Men | Women | Men | Women | Men | Women |
| Sodium [mg] | 3,142 | 2,200 | 2 | 16 | 1,500 | 1,500 |
| Chloride [mg] | 4,935 | 3,717 | 2 | 9 | 2,300 | 2,300 |
| Potassium [mg] | 2,508 | 2,325 | 97 | 98 | 4,000 | 4,000 |
| Calcium [mg] | 809 | 762 | 77 | 83 | 1,000 | 1,000 |
| Magnesium [mg] | 306 | 270 | 69 | 64 | 350 | 300 |
| Phosphorus [mg] | 1,233 | 1,009 | 0 | 2 | 550 | 550 |
| Zinc [mg] | 10.0 | 8.2 | 68 | 24 | 11* | 7* |
| Iron [mg] | 10.5 | 9.3 | 57 | 97 / 93<sup>2</sup> | 11 | 16 / 14<sup>2</sup> |
| Iodide [μg] | 80 | 71 | 99 | 100 | 200 | 200 |

<sup>1</sup> Reference values for 25–50-year-old men and women (33); <sup>2</sup> premenopausal/postmenopausal; \* for a diet with low phytate intake

### 5.5 Strengths and weaknesses

We present here for the first time after two decades detailed information on the diet of adults in Bavaria, describing the population distribution of food groups and nutrients. Employing the same methodology as in the previous BVS II enables a direct comparison with the dietary habits then and – hence – the description of the dietary changes over the past 20 years. We also applied the same food composition database in both studies (BLS 3.02), allowing a direct comparison between these studies. However, this can also be interpreted as a shortcoming, as an updated food composition database would have captured changes in food composition over time and included current food items, allowing for a more precise coding and nutrient calculation.

The field phase of the study took place during the SARS-CoV-2 pandemic. Besides potential effects on the response rate, which we addressed by weighting all analyses to compensate for any discrepancies with the underlying population, the pandemic may also have influenced dietary habits during the field phase, e.g., regarding out-of-home food consumption, particularly in the context of communal catering, which was not or not always possible at the time.

### 5.6 Conclusion

The present data describe changes in the dietary habits of the Bavarian adult population since the previous Bavarian Food Consumption Survey (BVS II) in 2002–2003.

The proportion of vegetarians and vegans has increased in the Bavarian population, and a flexitarian diet appears to become more prominent, resulting in a growing importance of meat alternatives and particularly milk alternatives in the daily diet, but also in a reduction in the consumption of red meat and especially processed meat. Other favorable changes compared to the BVS II include an increase in vegetable consumption, lower consumption of soft drinks, beer, and wine, as well as a concomitant increase in drinking water consumption.

However, a major proportion of the adult Bavarian population does not meet the current food-based dietary guidelines. Major deviations of the median intake from the FBDG of the DGE were observed for a wide range of important food groups, including fruit and vegetables, nuts and cereal products, particularly whole grain products. Accordingly, large proportions of the Bavarian population do not meet the DGE reference values for several essential nutrients, including folic acid, pantothenic acid, vitamin B6, iodine, calcium, and – previously not described – potassium.

The prevailing SARS-CoV-2 pandemic is likely to have affected the habitual diet. Subsequent surveys will have to examine the extent to which the situation has changed since the end of the pandemic.

## Ethics statement

At the beginning of the personal contact, detailed information about the aims and procedures of the study was provided and questions were answered; all participants gave written informed consent. The investigation was conducted according to the Declaration of Helsinki. All study methods involving human subjects were approved by the ethics review board at the Medical Faculty of the Ludwig-Maximilians-University of Munich (No. 20-0334).

## Author Contributions

FR: Data curation, Writing – original draft, Writing – review & editing. NW: Data curation, Formal Analysis, Methodology, Writing – review & editing. SG: Data curation, Writing – review & editing. NO: Writing – review & editing. MS: Project administration, Writing – review & editing. CR: Funding acquisition, Project administration, Writing – review & editing. MK: Project administration, Writing – review & editing. KG: Conceptualization, Funding acquisition, Resources, Writing – review & editing. JL: Conceptualization, Funding acquisition, Methodology, Resources, Supervision, Writing – original draft, Writing – review & editing.

## Acknowledgments

We are grateful to all colleagues who gave major input to the planning and conduct of the study. We thank the Max Rubner-Institute, Karlsruhe, Germany, for providing the current German version of the GloboDiet software. Special thanks go to the study participants as only through their commitment to participate and to provide data, this study could be conducted.

## Data Availability Statement

The study data will only be available after the publication of the basic results over the next months. Afterwards, the data are available upon justified request, i.e., providing a short description of the aims of the data analysis, and agreement of the project partners. In any case, misuse of the data has to be excluded.

