## Supplementary Tables for "Does the habitual dietary intake of adults in Bavaria, Germany, match dietary intake recommendations? – Results of the 3^rd^ Bavarian Food Consumption Survey (BVS III)"

**Supplementary Table S1: Food group consumption distribution (g/day) in male participants (n=550) of the BVS III, weighted for the deviation of the underlying population**

| **Food group,** subgroup | **Mean** | **SD** | **P5** | **P10** | **P25** | **P50** | **P75** | **P90** | **P95** |
| --- | --- | --- | --- | --- | --- | --- | --- | --- | --- |
| **Total meat** | 110.2 | 47.0 | 31.8 | 47.1 | 77.2 | 110.6 | 142.0 | 170.9 | 188.3 |
| **Fresh meat** | 61.5 | 22.1 | 26.5 | 32.7 | 44.9 | 60.8 | 77.1 | 91.2 | 98.8 |
| Red meat | 40.5 | 9.5 | 25.8 | 28.6 | 33.7 | 40.0 | 46.6 | 53.1 | 56.9 |
| Pork | 18.4 | 10.3 | 6.0 | 7.5 | 10.8 | 16.2 | 23.6 | 32.3 | 38.6 |
| Poultry | 20.6 | 11.6 | 8.0 | 9.6 | 13.1 | 18.1 | 24.5 | 33.0 | 41.7 |
| **Processed meat** | 48.9 | 27.1 | 11.2 | 16.0 | 27.6 | 45.6 | 66.4 | 85.9 | 98.0 |
| Sausages | 41.9 | 25.1 | 8.0 | 11.7 | 21.2 | 38.5 | 59.3 | 77.4 | 86.9 |
| Ham, cured meat | 7.4 | 5.8 | 1.4 | 1.9 | 3.3 | 5.8 | 9.8 | 15.1 | 19.2 |
| **Fish and seafood** | 18.0 | 17.2 | 2.5 | 3.7 | 6.8 | 12.7 | 23.0 | 38.7 | 51.5 |
| Fish, fresh/frozen | 9.3 | 7.7 | 1.9 | 2.6 | 4.3 | 7.3 | 11.9 | 18.2 | 23.5 |
| Fish, canned | 8.0 | 10.1 | 0.7 | 1.1 | 2.2 | 4.6 | 9.7 | 18.5 | 26.7 |
| **Eggs** | 18.7 | 14.2 | 3.2 | 4.7 | 8.4 | 15.1 | 25.3 | 37.8 | 46.5 |
| **Dairy products** | 140.0 | 79.8 | 38.0 | 51.7 | 82.1 | 125.3 | 182.5 | 245.0 | 289.9 |
| **Milk equivalents** | 433.8 | 218.1 | 131.4 | 177.8 | 275.8 | 404.3 | 561.2 | 724.6 | 835.9 |
| Milk | 62.1 | 70.3 | 4.0 | 7.0 | 17.0 | 38.9 | 81.5 | 145.3 | 198.0 |
| Cream | 0.9 | 2.3 | 0.0 | 0.0 | 0.1 | 0.3 | 0.8 | 2.1 | 3.7 |
| Fresh cheese, quark | 10.8 | 18.1 | 0.2 | 0.4 | 1.2 | 4.1 | 12.2 | 28.5 | 43.5 |
| Fermented dairy products (yogurt, kefir) | 25.3 | 32.4 | 0.8 | 1.5 | 4.1 | 12.2 | 32.9 | 69.2 | 96.2 |
| Other milk products (buttermilk, whey) | 6.5 | 26.4 | 0.0 | 0.0 | 0.0 | 0.3 | 1.8 | 10.5 | 28.3 |
| Cheese | 33.5 | 17.3 | 9.9 | 13.1 | 20.4 | 31.0 | 43.9 | 57.2 | 65.4 |
| **Butter** | 9.4 | 8.1 | 0.7 | 1.3 | 3.3 | 7.3 | 13.3 | 20.5 | 25.5 |
| **Other fats and oils** | 10.8 | 5.4 | 4.0 | 4.9 | 6.9 | 9.8 | 13.6 | 18.0 | 21.1 |
| Margarine | 1.4 | 3.9 | 0.0 | 0.0 | 0.1 | 0.3 | 1.1 | 3.5 | 6.5 |
| Vegetable oils | 4.6 | 1.9 | 1.7 | 2.2 | 3.1 | 4.4 | 5.9 | 7.2 | 8.0 |
| Mayonnaise | 4.5 | 3.8 | 0.7 | 1.0 | 1.8 | 3.4 | 6.0 | 9.4 | 12.1 |
| Fats and oils, unclassified | 1.0 | 0.2 | 0.8 | 0.8 | 0.8 | 0.9 | 1.1 | 1.2 | 1.2 |
| **Fat spreads (butter + margarine)** | 10.9 | 8.6 | 1.1 | 1.9 | 4.4 | 8.9 | 15.2 | 22.7 | 27.8 |
| **Cereal products** | 249.7 | 72.9 | 140.6 | 161.6 | 199.3 | 245.0 | 294.9 | 344.6 | 376.9 |
| **Bread and pastries** (non-whole grain) | 133.6 | 47.6 | 62.1 | 75.3 | 100.4 | 129.9 | 163.0 | 196.3 | 217.2 |
| White bread, crisp bread, rolls | 55.7 | 26.9 | 18.2 | 23.7 | 35.7 | 52.2 | 71.9 | 92.0 | 104.5 |
| Other types of bread (brown bread, spelt bread) | 27.2 | 22.8 | 3.2 | 5.1 | 10.4 | 21.1 | 37.3 | 57.3 | 72.1 |
| Pastries | 50.4 | 31.5 | 11.5 | 15.8 | 26.7 | 43.9 | 67.6 | 93.9 | 110.9 |
| **Pasta, rice, etc.** | 99.1 | 38.8 | 43.8 | 52.7 | 70.1 | 94.7 | 123.0 | 151.8 | 169.6 |
| Flour | 3.0 | 2.6 | 0.5 | 0.7 | 1.3 | 2.3 | 3.9 | 6.2 | 7.9 |
| Rice | 17.8 | 16.6 | 2.6 | 3.8 | 6.8 | 12.7 | 23.1 | 37.8 | 50.3 |
| Cereal grains (without rice) | 8.6 | 20.1 | 0.0 | 0.1 | 0.3 | 1.6 | 7.1 | 23.4 | 41.5 |
| Other cereal products | 4.2 | 5.5 | 0.3 | 0.5 | 1.0 | 2.3 | 5.1 | 10.1 | 14.7 |
| Pasta | 66.1 | 26.1 | 29.0 | 34.9 | 46.6 | 62.9 | 82.4 | 101.6 | 113.3 |
| **Whole grain products** | 17.0 | 13.0 | 2.7 | 3.9 | 7.2 | 13.5 | 23.7 | 35.4 | 43.1 |
| Muesli | 2.4 | 5.8 | 0.0 | 0.0 | 0.1 | 0.4 | 1.7 | 6.0 | 11.5 |
| Whole grain bread | 13.9 | 13.1 | 1.4 | 2.1 | 4.4 | 9.6 | 19.3 | 32.4 | 41.2 |
| **Potatoes** | 64.8 | 29.6 | 23.1 | 29.2 | 42.5 | 61.2 | 83.3 | 105.3 | 119.5 |
| Potatoes, fresh | 60.6 | 29.8 | 20.2 | 26.0 | 38.2 | 56.1 | 78.3 | 101.6 | 116.8 |
| **Fruit and vegetables** | 230.1 | 126.3 | 65.9 | 87.9 | 136.0 | 208.2 | 301.4 | 398.3 | 466.4 |
| **Vegetables** | 144.4 | 58.4 | 58.5 | 74.0 | 102.7 | 138.6 | 180.4 | 222.5 | 248.3 |
| Vegetables, unclassified | 14.2 | 9.5 | 3.2 | 4.3 | 7.1 | 11.9 | 18.9 | 27.4 | 33.4 |
| Salad | 18.4 | 11.8 | 4.5 | 6.1 | 9.7 | 15.7 | 24.2 | 34.3 | 41.1 |
| Leafy and stem vegetables | 5.2 | 7.5 | 0.4 | 0.6 | 1.3 | 2.8 | 6.1 | 11.7 | 17.5 |
| Cruciferous vegetables | 13.2 | 10.2 | 2.7 | 3.8 | 6.3 | 10.6 | 17.1 | 25.9 | 32.9 |
| Sprouting vegetables | 14.9 | 4.2 | 8.1 | 9.5 | 11.9 | 14.7 | 17.6 | 20.4 | 22.0 |
| Fruiting vegetables | 63.4 | 34.7 | 16.1 | 22.6 | 37.1 | 58.2 | 84.7 | 110.8 | 127.0 |
| Root vegetables | 12.8 | 6.2 | 5.0 | 6.0 | 8.3 | 11.6 | 15.9 | 20.8 | 24.4 |
| Mushrooms | 3.0 | 2.9 | 0.5 | 0.6 | 1.2 | 2.2 | 3.8 | 6.3 | 8.4 |
| Vegetable products | 2.0 | 4.8 | 0.0 | 0.0 | 0.1 | 0.4 | 1.6 | 4.8 | 8.9 |
| **Legumes** | 8.6 | 7.2 | 1.9 | 2.5 | 4.0 | 6.7 | 10.9 | 16.9 | 21.8 |
| **Fruit** | 86.1 | 74.7 | 5.1 | 9.7 | 27.2 | 67.4 | 124.9 | 189.2 | 232.4 |
| Fruit, unclassified | 6.0 | 11.8 | 0.1 | 0.2 | 0.7 | 2.0 | 5.8 | 14.9 | 25.3 |
| Pip fruits | 32.7 | 36.4 | 1.0 | 1.9 | 5.9 | 19.3 | 47.2 | 83.9 | 108.6 |
| Stone fruits | 6.5 | 17.6 | 0.0 | 0.1 | 0.2 | 0.9 | 4.1 | 15.3 | 31.3 |
| Berries | 6.6 | 12.8 | 0.1 | 0.2 | 0.6 | 2.0 | 6.4 | 17.4 | 29.1 |
| Tropical fruits | 27.9 | 38.9 | 0.5 | 1.0 | 3.5 | 12.7 | 36.9 | 75.0 | 106.2 |
| Citrus fruits | 9.8 | 26.7 | 0.0 | 0.0 | 0.3 | 1.4 | 6.8 | 23.9 | 46.9 |
| **Nuts** | 7.1 | 10.2 | 0.3 | 0.6 | 1.3 | 3.3 | 8.4 | 18.4 | 27.0 |
| **Sugar** | 1.0 | 1.8 | 0.0 | 0.0 | 0.1 | 0.4 | 1.1 | 2.5 | 3.9 |
| **Jam** | 5.1 | 6.7 | 0.0 | 0.1 | 0.4 | 2.1 | 7.6 | 15.0 | 19.4 |
| **Sweets** | 17.0 | 15.5 | 2.1 | 3.2 | 6.3 | 12.5 | 22.8 | 36.9 | 47.1 |
| Chocolate and chocolate products | 6.7 | 7.7 | 0.6 | 0.9 | 1.8 | 4.0 | 8.7 | 16.2 | 22.1 |
| Confectioneries and other sweets | 1.6 | 3.4 | 0.0 | 0.1 | 0.2 | 0.5 | 1.5 | 3.9 | 6.9 |
| Ice cream | 5.0 | 9.3 | 0.1 | 0.2 | 0.6 | 1.7 | 5.0 | 12.8 | 21.3 |
| Honey | 3.2 | 5.1 | 0.1 | 0.1 | 0.4 | 1.3 | 3.9 | 8.9 | 13.1 |
| **Desserts** | 13.2 | 4.4 | 6.5 | 7.8 | 10.4 | 13.0 | 15.5 | 19.1 | 21.5 |
| **Soups and sauces** | 43.2 | 23.5 | 14.6 | 18.2 | 26.2 | 38.5 | 55.0 | 74.3 | 87.5 |
| **Spices and condiments** | 18.1 | 8.8 | 5.7 | 7.5 | 11.5 | 16.9 | 23.4 | 30.0 | 34.3 |
| **Alternatives** | 12.1 | 33.0 | 0.1 | 0.1 | 0.4 | 1.2 | 5.0 | 31.2 | 79.3 |
| Milk alternatives | 10.5 | 33.8 | 0.0 | 0.0 | 0.1 | 0.3 | 2.0 | 19.6 | 75.2 |
| Meat alternatives | 2.2 | 6.7 | 0.0 | 0.1 | 0.1 | 0.4 | 1.2 | 4.1 | 10.0 |
| **Non-alcoholic beverages** | 1764.0 | 815.0 | 564.9 | 764.2 | 1168.8 | 1688.2 | 2280.1 | 2857.3 | 3231.6 |
| Juices | 33.3 | 66.0 | 0.4 | 0.9 | 3.1 | 10.5 | 33.5 | 85.6 | 142.1 |
| Water | 1511.4 | 860.7 | 172.1 | 384.5 | 875.3 | 1459.5 | 2072.7 | 2644.5 | 3014.5 |
| Spritzers | 69.0 | 131.2 | 0.1 | 0.4 | 1.9 | 11.3 | 62.8 | 227.5 | 382.2 |
| Soft drinks | 139.1 | 195.8 | 0.8 | 1.8 | 7.5 | 40.3 | 201.4 | 451.0 | 571.5 |
| Other non-alcoholic beverages | 26.3 | 57.1 | 0.4 | 0.7 | 2.2 | 7.2 | 23.3 | 66.1 | 116.3 |
| Coffee substitutes | 20.6 | 43.4 | 0.3 | 0.6 | 1.7 | 5.7 | 18.8 | 51.8 | 92.8 |
| **Coffee** | 285.8 | 234.7 | 7.3 | 21.2 | 97.4 | 246.8 | 414.0 | 598.6 | 731.3 |
| **Tea and other infusions** | 150.0 | 267.1 | 0.0 | 0.2 | 1.7 | 22.1 | 183.4 | 491.8 | 704.9 |
| Tea | 70.3 | 152.0 | 0.1 | 0.2 | 1.1 | 8.1 | 58.3 | 222.9 | 379.1 |
| Other infusions | 81.6 | 183.7 | 0.1 | 0.2 | 1.3 | 9.4 | 64.6 | 248.0 | 440.7 |
| **Alcoholic beverages** | 236.1 | 281.7 | 2.9 | 7.3 | 29.8 | 123.7 | 347.7 | 639.7 | 830.5 |
| Beer | 202.6 | 262.7 | 2.2 | 5.2 | 21.0 | 90.1 | 290.6 | 568.7 | 758.2 |
| Wine, champagne | 28.4 | 59.2 | 0.1 | 0.3 | 1.3 | 6.2 | 25.5 | 78.2 | 139.1 |

SD: standard deviation, P: percentile

**Supplementary Table S2: Food group consumption distributions (g/day) in female participants (n=689) of the BVS III, weighted for the deviation of the underlying population**

| **Food group,** subgroup | **Mean** | **SD** | **P5** | **P10** | **P25** | **P50** | **P75** | **P90** | **P95** |
| --- | --- | --- | --- | --- | --- | --- | --- | --- | --- |
| **Total meat** | 71.9 | 39.2 | 13.4 | 21.4 | 41.0 | 69.6 | 98.7 | 124.6 | 140.2 |
| **Fresh meat** | 41.4 | 17.9 | 15.8 | 19.5 | 27.6 | 39.4 | 53.2 | 66.4 | 74.3 |
| Red meat | 25.0 | 7.1 | 14.9 | 16.6 | 19.9 | 24.1 | 29.1 | 34.3 | 37.8 |
| Pork | 10.5 | 6.7 | 3.2 | 3.9 | 5.8 | 8.8 | 13.4 | 19.3 | 23.6 |
| Poultry | 16.3 | 9.8 | 5.9 | 7.0 | 9.6 | 13.7 | 20.0 | 28.6 | 37.2 |
| **Processed meat** | 29.9 | 20.0 | 5.7 | 8.0 | 14.4 | 25.8 | 41.2 | 57.9 | 68.1 |
| Sausages | 22.6 | 16.8 | 3.5 | 5.0 | 9.4 | 18.0 | 32.0 | 47.2 | 55.7 |
| Ham, cured meat | 6.9 | 5.4 | 1.3 | 1.8 | 3.0 | 5.3 | 9.0 | 14.0 | 17.6 |
| **Fish and seafood** | 13.3 | 12.9 | 1.8 | 2.6 | 4.8 | 9.2 | 17.3 | 28.8 | 38.5 |
| Fish, fresh/frozen | 7.8 | 6.3 | 1.5 | 2.1 | 3.5 | 6.0 | 10.1 | 15.5 | 19.9 |
| Fish, canned | 4.3 | 5.6 | 0.4 | 0.6 | 1.1 | 2.4 | 5.3 | 10.0 | 14.5 |
| **Eggs** | 18.8 | 13.6 | 3.4 | 4.9 | 8.8 | 15.6 | 25.4 | 37.2 | 45.3 |
| **Dairy products** | 159.6 | 85.2 | 49.6 | 65.2 | 97.9 | 144.2 | 205.5 | 273.5 | 319.3 |
| **Milk equivalents** | 459.5 | 216.1 | 161.0 | 208.3 | 301.9 | 428.9 | 586.0 | 751.4 | 855.9 |
| Milk | 66.3 | 71.2 | 5.1 | 8.8 | 19.6 | 43.6 | 87.5 | 152.1 | 203.0 |
| Cream | 1.8 | 3.9 | 0.0 | 0.1 | 0.2 | 0.6 | 1.7 | 4.3 | 7.3 |
| Fresh cheese, quark | 14.4 | 22.0 | 0.3 | 0.6 | 1.9 | 6.3 | 17.6 | 37.6 | 55.1 |
| Fermented dairy products (yogurt, kefir) | 40.8 | 40.0 | 2.0 | 3.7 | 9.6 | 26.6 | 61.2 | 100.3 | 123.9 |
| Other milk products (buttermilk, whey) | 4.8 | 19.9 | 0.0 | 0.0 | 0.0 | 0.2 | 1.4 | 8.0 | 21.0 |
| Cheese | 31.9 | 15.7 | 10.1 | 13.2 | 20.1 | 29.7 | 41.5 | 53.3 | 60.6 |
| **Butter** | 7.4 | 6.5 | 0.5 | 1.0 | 2.5 | 5.7 | 10.6 | 16.3 | 20.3 |
| **Other fats and oils** | 10.1 | 5.1 | 3.7 | 4.5 | 6.4 | 9.1 | 12.8 | 16.9 | 19.8 |
| Margarine | 1.2 | 3.2 | 0.0 | 0.0 | 0.1 | 0.2 | 1.0 | 3.1 | 5.7 |
| Vegetable oils | 5.4 | 2.1 | 2.1 | 2.6 | 3.8 | 5.3 | 6.9 | 8.2 | 8.9 |
| Mayonnaise | 2.3 | 2.1 | 0.3 | 0.5 | 0.9 | 1.7 | 3.1 | 5.0 | 6.4 |
| Fats and oils, unclassified | 0.5 | 0.1 | 0.4 | 0.4 | 0.5 | 0.5 | 0.6 | 0.7 | 0.7 |
| **Fat spreads (butter + margarine)** | 8.6 | 7.0 | 0.8 | 1.5 | 3.3 | 6.9 | 12.1 | 18.1 | 22.2 |
| **Cereal products** | 193.8 | 62.6 | 101.4 | 118.5 | 150.0 | 188.2 | 232.4 | 276.3 | 304.1 |
| **Bread and pastries** (non-whole grain) | 92.0 | 37.5 | 37.4 | 46.8 | 65.0 | 88.2 | 114.8 | 141.9 | 159.9 |
| White bread, crisp bread, rolls | 32.5 | 19.0 | 8.5 | 11.5 | 18.3 | 28.9 | 43.0 | 58.6 | 68.8 |
| Other types of bread (brown bread, spelt bread) | 18.8 | 16.0 | 2.2 | 3.4 | 6.9 | 14.3 | 26.0 | 40.2 | 50.2 |
| Pastries | 40.8 | 26.0 | 9.1 | 12.6 | 21.1 | 35.5 | 54.9 | 76.6 | 90.8 |
| **Pasta, rice, etc.** | 85.8 | 34.2 | 37.7 | 45.1 | 60.4 | 81.6 | 106.7 | 132.0 | 148.1 |
| Flour | 3.3 | 2.9 | 0.5 | 0.8 | 1.4 | 2.5 | 4.3 | 6.8 | 8.8 |
| Rice | 19.6 | 17.7 | 3.0 | 4.2 | 7.5 | 14.1 | 25.7 | 41.8 | 54.9 |
| Cereal grains (without rice) | 10.5 | 21.3 | 0.1 | 0.1 | 0.5 | 2.5 | 10.6 | 30.2 | 48.3 |
| Other cereal products | 4.3 | 5.4 | 0.3 | 0.5 | 1.0 | 2.4 | 5.3 | 10.3 | 14.9 |
| Pasta | 47.1 | 20.1 | 19.9 | 24.0 | 32.2 | 44.1 | 58.9 | 74.3 | 84.5 |
| **Whole grain products** | 15.7 | 11.2 | 2.7 | 3.9 | 7.1 | 13.1 | 21.7 | 31.5 | 38.0 |
| Muesli | 3.5 | 7.5 | 0.0 | 0.0 | 0.2 | 0.7 | 2.9 | 9.4 | 17.6 |
| Whole grain bread | 12.3 | 10.8 | 1.3 | 2.0 | 4.1 | 9.0 | 17.4 | 27.6 | 34.2 |
| **Potatoes** | 58.6 | 27.0 | 20.8 | 26.3 | 37.9 | 55.3 | 75.9 | 95.6 | 107.8 |
| Potatoes, fresh | 55.0 | 27.2 | 18.3 | 23.4 | 34.2 | 51.0 | 71.7 | 92.4 | 105.6 |
| **Fruit and vegetables** | 309.4 | 154.3 | 103.2 | 132.9 | 196.7 | 285.7 | 395.1 | 517.2 | 600.1 |
| **Vegetables** | 171.0 | 64.6 | 75.4 | 92.4 | 124.9 | 165.4 | 210.7 | 256.4 | 286.4 |
| Vegetables, unclassified | 10.3 | 7.4 | 2.1 | 2.9 | 4.9 | 8.4 | 13.9 | 20.6 | 25.2 |
| Salad | 17.3 | 11.2 | 4.2 | 5.7 | 9.1 | 14.8 | 22.9 | 32.3 | 38.9 |
| Leafy and stem vegetables | 5.2 | 7.8 | 0.3 | 0.6 | 1.2 | 2.7 | 6.0 | 11.9 | 17.9 |
| Cruciferous vegetables | 15.9 | 11.9 | 3.5 | 4.7 | 7.7 | 12.8 | 20.6 | 30.9 | 38.7 |
| Sprouting vegetables | 15.3 | 4.1 | 8.7 | 10.1 | 12.5 | 15.2 | 17.9 | 20.6 | 22.3 |
| Fruiting vegetables | 81.1 | 39.7 | 24.1 | 32.4 | 50.8 | 77.2 | 105.9 | 134.5 | 152.3 |
| Root vegetables | 15.4 | 7.4 | 6.2 | 7.5 | 10.2 | 13.9 | 19.2 | 25.1 | 29.3 |
| Mushrooms | 3.0 | 2.9 | 0.4 | 0.6 | 1.1 | 2.1 | 3.9 | 6.4 | 8.4 |
| Vegetable products | 3.2 | 7.0 | 0.0 | 0.1 | 0.2 | 0.8 | 2.8 | 8.1 | 14.7 |
| **Legumes** | 11.4 | 9.1 | 2.6 | 3.4 | 5.5 | 8.9 | 14.5 | 22.2 | 28.6 |
| **Fruit** | 133.1 | 92.6 | 15.4 | 27.8 | 61.7 | 117.1 | 185.7 | 260.0 | 308.9 |
| Fruit, unclassified | 9.9 | 17.2 | 0.2 | 0.4 | 1.2 | 3.5 | 10.6 | 25.9 | 42.0 |
| Pip fruits | 39.7 | 37.6 | 1.7 | 3.5 | 10.2 | 28.1 | 59.0 | 93.7 | 116.1 |
| Stone fruits | 9.2 | 18.5 | 0.1 | 0.1 | 0.4 | 1.9 | 8.3 | 26.4 | 46.4 |
| Berries | 15.0 | 21.3 | 0.4 | 0.7 | 2.0 | 6.4 | 18.9 | 41.8 | 60.0 |
| Tropical fruits | 38.9 | 46.0 | 1.2 | 2.3 | 7.1 | 22.2 | 54.1 | 98.6 | 131.3 |
| Citrus fruits | 16.8 | 37.5 | 0.0 | 0.1 | 0.6 | 3.3 | 14.4 | 45.5 | 80.4 |
| **Nuts** | 7.0 | 9.8 | 0.3 | 0.5 | 1.2 | 3.3 | 8.6 | 18.0 | 26.2 |
| **Sugar** | 1.3 | 2.3 | 0.0 | 0.1 | 0.2 | 0.5 | 1.5 | 3.3 | 5.2 |
| **Jam** | 5.1 | 6.4 | 0.0 | 0.1 | 0.4 | 2.2 | 7.8 | 14.4 | 18.5 |
| **Sweets** | 18.2 | 15.8 | 2.4 | 3.6 | 7.1 | 13.7 | 24.5 | 38.3 | 48.9 |
| Chocolate and chocolate products | 7.6 | 8.1 | 0.6 | 1.0 | 2.0 | 4.8 | 10.2 | 17.9 | 23.7 |
| Confectioneries and other sweets | 1.6 | 3.3 | 0.0 | 0.1 | 0.2 | 0.5 | 1.5 | 4.0 | 6.8 |
| Ice cream | 6.3 | 10.3 | 0.2 | 0.3 | 0.8 | 2.4 | 7.0 | 16.7 | 26.2 |
| Honey | 2.5 | 3.7 | 0.0 | 0.1 | 0.3 | 1.0 | 3.1 | 6.8 | 9.9 |
| **Desserts** | 13.0 | 3.9 | 7.4 | 8.3 | 10.0 | 12.9 | 15.2 | 17.5 | 20.2 |
| **Soups and sauces** | 33.2 | 18.6 | 10.9 | 13.6 | 19.7 | 29.2 | 42.8 | 57.9 | 68.6 |
| **Spices and condiments** | 17.2 | 8.3 | 5.6 | 7.3 | 10.9 | 16.2 | 22.3 | 28.5 | 32.3 |
| **Alternatives** | 13.3 | 29.1 | 0.1 | 0.2 | 0.5 | 1.8 | 8.5 | 43.5 | 78.2 |
| Milk alternatives | 11.1 | 29.5 | 0.0 | 0.0 | 0.1 | 0.4 | 3.5 | 35.4 | 74.5 |
| Meat alternatives | 3.2 | 8.3 | 0.0 | 0.1 | 0.2 | 0.6 | 2.1 | 7.6 | 16.1 |
| **Non-alcoholic beverages** | 1617.4 | 788.6 | 477.6 | 660.8 | 1033.4 | 1532.9 | 2108.4 | 2681.0 | 3034.2 |
| Juices | 29.1 | 58.8 | 0.3 | 0.7 | 2.3 | 8.5 | 28.5 | 76.9 | 128.2 |
| Water | 1401.3 | 795.9 | 200.6 | 398.2 | 806.1 | 1335.0 | 1919.1 | 2468.5 | 2818.3 |
| Spritzers | 45.4 | 92.9 | 0.1 | 0.2 | 1.0 | 6.3 | 36.7 | 146.1 | 254.6 |
| Soft drinks | 62.5 | 110.8 | 0.3 | 0.7 | 2.8 | 13.5 | 64.2 | 201.5 | 318.8 |
| Other non-alcoholic beverages | 20.2 | 42.4 | 0.3 | 0.5 | 1.6 | 5.6 | 18.8 | 51.8 | 89.9 |
| Coffee substitutes | 25.9 | 53.0 | 0.4 | 0.7 | 2.1 | 7.2 | 24.9 | 65.9 | 115.1 |
| **Coffee** | 311.5 | 237.7 | 14.0 | 35.8 | 127.7 | 273.4 | 443.4 | 623.3 | 757.6 |
| **Tea and other infusions** | 296.9 | 396.5 | 0.2 | 1.0 | 11.5 | 123.7 | 460.2 | 828.5 | 1084.8 |
| Tea | 106.7 | 212.1 | 0.1 | 0.3 | 1.8 | 14.6 | 103.2 | 349.7 | 537.5 |
| Other infusions | 190.2 | 303.7 | 0.4 | 1.2 | 7.3 | 53.1 | 253.6 | 569.3 | 805.2 |
| **Alcoholic beverages** | 92.7 | 143.9 | 0.5 | 1.3 | 6.0 | 30.2 | 116.8 | 274.6 | 396.7 |
| Beer | 50.0 | 101.2 | 0.2 | 0.5 | 2.0 | 10.0 | 46.4 | 145.4 | 252.0 |
| Wine, champagne | 40.2 | 70.3 | 0.3 | 0.6 | 2.5 | 11.4 | 44.0 | 116.3 | 185.7 |

SD: standard deviation, P: percentile
